# The Audible Contrast Threshold (ACT™) test: a clinical spectro-temporal modulation detection test

**DOI:** 10.1101/2023.10.12.23296977

**Authors:** Johannes Zaar, Lisbeth Birkelund Simonsen, Raul Sanchez-Lopez, Søren Laugesen

## Abstract

Over the last decade, multiple studies have shown that hearing-impaired listeners’ speech-in-noise reception ability, measured with audibility compensation, is closely associated with performance in spectro-temporal modulation (STM) detection tests. STM tests thus have the potential to provide highly relevant beyond-the-audiogram information in the clinic, but the available STM tests have not been optimized for clinical use in terms of test duration, required equipment, and procedural standardization. The present study introduces a quick-and-simple clinically viable STM test, named the Audible Contrast Threshold (ACT) test. First, an experimenter-controlled STM measurement paradigm was developed, in which the patient is presented binaurally with a continuous audibility-corrected noise via headphones and asked to press a pushbutton whenever they hear an STM target sound in the noise. The patient’s threshold is established using a Hughson-Westlake tracking procedure with a three-out-of-five criterion and then refined by post-processing the collected data using a logistic function. Different stimulation paradigms were tested in 28 hearing-impaired participants and compared to data previously measured in the same participants with an established STM test paradigm. The best stimulation paradigm showed excellent test-retest reliability and good agreement with the established laboratory version. Second, the best stimulation paradigm with 1-second noise “waves” (windowed noise) was chosen, further optimized with respect to step size and logistic-function fitting, and tested in a population of 25 young normal-hearing participants using various types of transducers to obtain normative data. Based on these normative data, the “normalized Contrast Level” (in dB nCL) scale was defined, where 0±4 dB nCL corresponds to normal performance and the greater the positive value of dB nCL, the greater the audible contrast loss. Overall, the results of the present study indicate that the ACT test may be considered a reliable, quick-and-simple (and thus clinically viable) test of STM sensitivity. The ACT can be measured directly after the audiogram using the same set up, adding only a few minutes to the process.

**CITE AS:**

1) In text: Zaar/Simonsen et al. (2023)
2) In reference list: Zaar, J./Simonsen L. B., Sanchez-Lopez, R., and Laugesen, S. (2023): “The Audible Contrast Threshold (ACT^™^) test: a clinical spectro-temporal modulation detection test,” medRxiv.

## Introduction

Pure-tone audiometry is one of the first diagnostic measurements that adult hearing-impaired (HI) individuals are exposed to when in need of hearing aids. The audiogram determines the degree of amplification to be applied in various frequency regions to ensure audibility when fitting a hearing aid. However, individuals with a sensorineural hearing loss often experience great difficulties in understanding speech in noisy environments (Kochkin, 2000), also referred to as “supra-threshold distortion”, which is not necessarily well predicted from pure-tone audiometry (Vermiglio and Fang, 2022) and which amplification alone does not solve (Plomp, 1978; Grant et al., 2013). Most of today’s state-of-the-art hearing aids have the ability to improve speech understanding in challenging speech-in-noise situations, but the need for optimal fitting of these sound-processing features vary greatly among individuals (Andersen et al., 2021; Zaar et al., 2023b). To fit the hearing aids such that each patient’s individual needs are met, the clinician needs information about the degree to which the patient is struggling in real life speech-in-noise scenarios, when audibility is compensated for. Currently, the individualized fitting of advanced hearing-aid parameters, such as the degree of noise reduction, is typically estimated by the clinician based on conversations with the patient, patient characteristics (e.g., age, lifestyle) and intuition, or based on the conventional speech audiometry in quiet (ISO 8253-3, 2012). However, a number of studies suggest poor predictive power of the speech-in-quiet measures with respect to any speech-in-noise measure (Duquesnoy, 1983; Grant and Walden, 2013; Killion et al., 2004; Killion and Niquette, 2000; Nilsson et al., 1994; Smoorenburg, 1992). Thus, speech-in-quiet tests have limited utility for the individualization of the hearing-aid fitting.

Given the above considerations, the “ecological validity” of hearing tests should be taken into account, referring to the extent to which the results of an evaluation are representative of real-world auditory ability (Keidser et al., 2020). An ecologically valid speech-in-noise test should thus be carried out while wearing hearing aids and ideally involve multi-talker babble presented from spatially separated loudspeakers. This setup has been shown in literature (Neher et al., 2009; Rønne et al., 2017; Zaar et al., 2023a, 2023b) to establish an individual’s speech-in-noise performance more precisely than, e.g., an unaided speech-in-noise test with steady-state noise presented via headphones (Nilsson et al., 1994) or from a single loudspeaker (Killion et al., 2004), even with individualized amplification provided (Zaar et al., 2023a). However, performing an ecologically valid speech-in-noise test is complex and time-consuming, such that it may not be a viable option for clinical purposes (Parmar et al., 2022), whereas simple clinical speech-in-noise tests have been shown to yield limited separation between relevant populations (Billings et al., 2023).

In addition to the missing link between speech-in-*noise* performance and speech-in-*quiet* measurements, all speech tests are necessarily language dependent and thus require that the speech material match the native language of the patient to avoid language proficiency bias. Moreover, speech-test results may be biased by learning effects (Hällgren et al., 2006; Hernvig and Olsen, 2005; Nielsen and Dau, 2011; Simonsen et al., 2016; Yund and Woods, 2010) as the available validated speech material is limited in various languages. According to a recent study (Parmar et al., 2022), the main reason for not carrying out speech-in-noise tests in the clinic was time limitations; hearing health care professionals furthermore identified “lack of speech test material in non-English language”, “lack of normative data”, and “no clear evidence for patient benefit or care” as significant barriers to a wider clinical use of speech tests.

Clinicians are thus in need of a diagnostic test that allows to optimize the individualization of advanced hearing-aid fitting parameters based on the patient’s supra-threshold hearing deficits.

Such a test should:

- Predict the individual patient’s speech-in-noise performance measured in an ecologically valid speech test with individualized hearing-aid amplification.
- Be quick and simple (similar to conventional speech-in-quiet measurements, and different from complex and time-consuming ecologically valid speech-in-noise tests).
- Be language independent.

An excellent candidate for such a diagnostic test is the spectro-temporal modulation (STM) detection test, a psychoacoustic test that estimates the degree of modulation required for an individual to discriminate the modulated target from an unmodulated reference stimulus. The target is a broadband noise carrier that is modulated both spectrally and temporally, thus creating spectral ripples that move up or down in frequency as a function of time, while the reference sound is the noise carrier without modulation. Different types of STM tests have been investigated with normal-hearing (NH) as well as HI individuals and were found to be promising, as several studies demonstrated that the STM detection thresholds predicted speech-in-noise performance (Bernstein et al., 2016, 2013; Mehraei et al., 2014; Zaar et al., 2023a, 2023b). These studies revealed that the prediction of speech-in-noise performance is more robust when the STM detection thresholds are combined with speech-audibility prediction using the speech intelligibility index (ANSI S3.5, 1997; Bernstein et al., 2013; Mehraei et al., 2014) or with the pure-tone thresholds directly (Bernstein et al., 2016; Zaar et al., 2023b) and furthermore indicated a benefit when the STM stimuli are sufficiently audible (Zaar et al., 2023a, 2023b).

The STM stimuli are language independent, but they mimic the essential spectral and temporal modulations contained in the speech signal. Therefore, it is reasonable to expect that a reduction in the ability to detect STM is associated with a reduced speech understanding. Overall, the supra-threshold distortions of HI individuals are typically ascribed to the impairment of three essential mechanisms of the auditory system, namely a reduced ability to use temporal fine-structure information, impaired temporal resolution, and reduced spectral resolution; all three mechanisms are employed in an STM detection task, which may (partly) explain the strong relationship to speech-in-noise outcomes (Mehraei et al., 2014). Furthermore, the detection of the STM stimuli can be connected to supra-threshold sensitivity to acoustic contrasts (Lunner et al., 2020). The hearing abilities that allow the patient to identify subtle differences in the STM stimuli and perceive complex spectro-temporal patterns may therefore be connected to their ability to extract relevant target-speech features in noisy environments. Thus, in analogy to assessing hearing sensitivity with respect to sound level by way of pure-tone audiometry, the STM detection can be understood as a measure of audible contrast sensitivity.

In summary, STM sensitivity has been established as a good predictor of individual speech-in-noise performance and is language independent by design. One question remains: is the measurement of STM sensitivity also quick and simple? The STM tests used in previous studies were, in fact, rather time-consuming due to the need for extensive training of about 1 hour in some studies (Bernstein et al., 2013; Mehraei et al., 2014), and an effective testing duration of at least 15 minutes (Bernstein et al., 2016; Zaar et al., 2023a, 2023b). It should be noted that the mentioned studies applied high-precision research procedures that were neither optimized nor intended for clinical use. These psychoacoustic procedures were based either on a two-alternative forced-choice (2-AFC) task (Bernstein et al., 2016, 2013; Mehraei et al., 2014) or on a 3-AFC task (Zaar et al., 2023a, 2023b), in which the test participants listened to a sequence of two or three sounds, whereafter the task was to choose the sound that contained the modulation.

Alternative STM detection tests have been proposed using simpler tasks (Yes/No task; Sanchez-Lopez et al., 2021; 4-interval 2-AFC; Gallun et al., 2022). However, the reduction in testing time and complexity of these alternative test paradigms might also have affected the predictive power of the results compared to the previous approaches (Gallun et al., 2022).

Therefore, the above-mentioned STM test paradigms do not meet the need of being quick and simple, which thus remains as the main requirement to be met for clinical viability. To develop a clinical diagnostic test, the threshold-seeking procedure should be easy to administer as, for example, the modified Hughson-Westlake method (Hughson and Westlake, 1944; Carhart and Jerger, 1959) used in pure-tone audiometry. Additionally, the test results may be presented as the deviation in performance from young NH listeners as a reference. This would allow the clinician to quantify the results based on the supra-threshold hearing abilities (contrast sensitivity) rather than in terms of the properties of the physical stimulus (modulation level). The present article documents the development of a clinical test for assessing supra-threshold distortions: the Audible Contrast Threshold (ACT) test. The ACT represents the minimum contrast required by the patient to detect an STM stimulus that is sufficiently audible. Two studies are presented that together establish a quick-and-simple STM test in combination with a normative performance range. Study A explores different ways of implementing a quick-and-simple clinical STM measurement approach without compromising its reliability and predictive connection to speech-in-noise outcomes, evaluated by re-testing the HI test participants from Zaar et al. (2023b) and using their 3-AFC-based STM scores as a reference. Special attention was paid to the clinical viability with respect to using only headphones (or insert phones) and pushbutton and employing a test paradigm that was strongly inspired by the audiogram measurement approach. Study B presents a matured version of the selected measurement approach implemented as a research prototype software and establishes normative data by testing 25 young NH test participants. In analogy to the definition of audiometric thresholds in terms of hearing level (HL) based on the Reference Equivalent Threshold Sound Pressure Level (RETSPL), the normative data from NH listeners here represents the Equivalent Threshold Modulation Level (ETML), which is used to establish the zero-reference in terms of normative Contrast Level (nCL) for different transducers.

## Study A: Developing a quick and simple clinical STM detection test

### Background

Zaar et al. (2023b) measured ecologically valid speech reception thresholds (SRTs) in a speech-in-noise test setup with spatialized multi-talker interferers for 30 HI participants equipped with hearing aids. In addition, STM detection thresholds were obtained using a 3-AFC measurement paradigm for two different STM stimulus variants, which were selected based on a previous exploratory study (Zaar et al., 2023a). The “Noisy-LP_2co_” variant employing a bandlimited noise carrier (354-2000 Hz), a spectral modulation frequency of 2 cycles/octave (c/o), and a temporal modulation frequency of 4 Hz (with upward moving ripples) was selected for further development due to far superior test-retest reliability as compared to a stimulus with a complex-tone carrier (“Tonal_2co_”). The STM detection thresholds were found to be strongly correlated with the measured SRTs (R^2^=0.61) as well as with the speech-intelligibility benefit induced by strong (compared to mild) hearing-aid directionality and noise reduction processing (R^2^=0.51). Furthermore, the data collected by Zaar et al. (2023b) showed a non-significant potential connection between poor STM performance and preference for strong directionality and noise reduction hearing-aid processing in the field. Thus, the STM detection threshold shows strong potential for clinical assessment of supra-threshold hearing deficits and potentially for prescription of hearing-aid signal processing. However, the measurement paradigm employed by Zaar et al. (2023a, 2023b) was not optimized for clinical practice as it requires a computer interface and monitor for the test person to conduct the test and is rather time consuming (about 15 minutes).

Study A has therefore been conducted as a follow-up to the Zaar et al. (2023b) study, testing the same test participants with two different measurement paradigms designed for clinical practice with the goal that they (i) are quick, (ii) can be conducted with only a pair of headphones and a pushbutton, and (iii) bear maximum resemblance with the measurement method applied in pure-tone audiometry (Carhart and Jerger, 1959). The two measurement paradigms were tested with two different step sizes, amounting to four test conditions. To improve threshold estimation accuracy, a suitable post-processing strategy of the collected data using a logistic function fit was applied. The different test conditions were compared with respect to group performance level, measurement duration, test-retest reliability, and agreement with the reference STM thresholds originally measured by Zaar et al. (2023b) using the mentioned 3-AFC procedure.

## Methods

### Test participants

The 30 test participants from Zaar et al. (2023b) were re-invited to participate in the present study, which was conducted approximately one year after the initial visit of the Zaar et al. (2023b) study. Twenty-eight of the original 30 participants were successfully recruited (9 female). The age of the participants ranged from 46 to 82 years (mean: 71.2 years, standard deviation: 9.4 years). All participants were regular hearing-aid users. Their audiograms are illustrated in Figure 1, showing a range of mild to severe/profound hearing losses. The individual audiograms were largely symmetric, as 27 participants showed thresholds within 15 dB between ears for at least 9 out of the 11 test frequencies and one participant for 7 of the 11 test frequencies. Test participants provided informed consent and experiments were approved by the Science-Ethics Committee for the Capital Region of Denmark (Study A, reference H-16036391).

**Figure 1:**
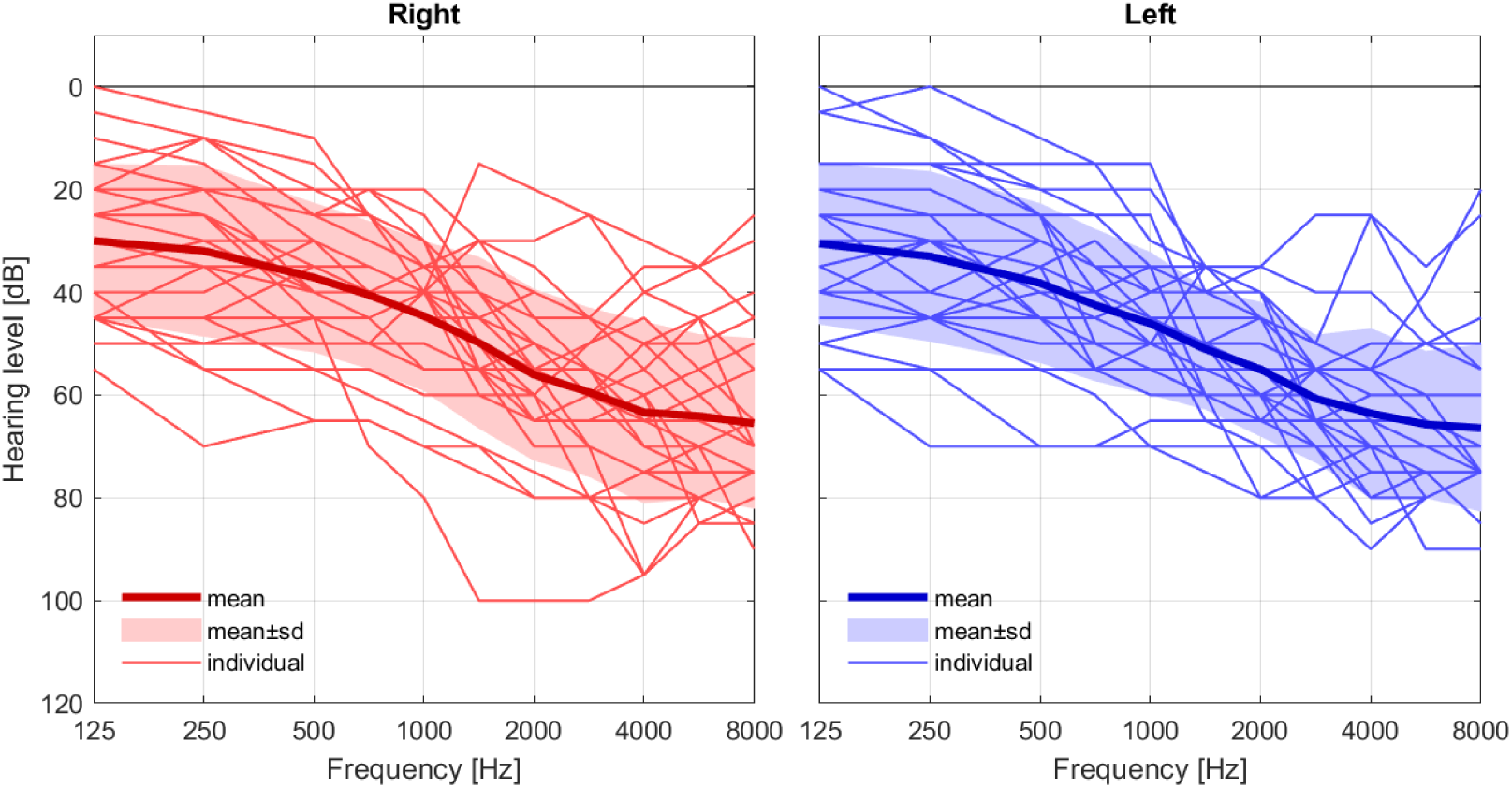
Audiograms of the study-A test participants. The thin lines show the audiograms for individual participants, the thick lines the across-participant mean, and the shaded areas reflect the across-participant standard deviations.

### Stimulation and measurement paradigms

All stimuli were based on the Noisy-LP_2co_ variant from Zaar et al. (2023a, 2023b), which consists of a pink-noise carrier signal in the band from 354 to 2000 Hz and an upward-moving STM with a temporal modulation rate of 4 Hz and a spectral modulation frequency of 2 c/o. The noise carrier was created as a sum of 2499 sinusoidal components with random starting phase, which were logarithmically distributed along the frequency axis in the considered band. For each presentation, the starting phase of the STM pattern was randomly selected from a uniform distribution in the range [−*π*, *π*]. Mathematical details on the stimulus generation are found in Zaar et al. (2023a).

The stimulation paradigm was inspired by pure-tone audiometry. During the test, a constant carrier noise was used as the reference, while an STM (target) imposed on the carrier was played by pressing the “stimulus” button (similar to the way pure tones are presented in an audiometry), see also Table 1. The test participant was asked to press a pushbutton when they heard the modulation. Two paradigms were considered:

1. *On-going*, where the noise carrier was played continuously. A 1-s long STM was imposed when the examiner pushed the stimulus button (see top panel of Figure 2 for an illustration).
2. *Waves*, where 1-second-long carrier waves, generated by imposing 125-ms long raised-cosine fade in/outs onto 1-s long carrier signals, were concatenated such that there was a 250-ms long fading out and fading back in portion in between two consecutive waves. The STM was imposed on the next carrier wave when the examiner pushed the stimulus button (see bottom panel of Figure 2 for an illustration).

**Figure 2:**
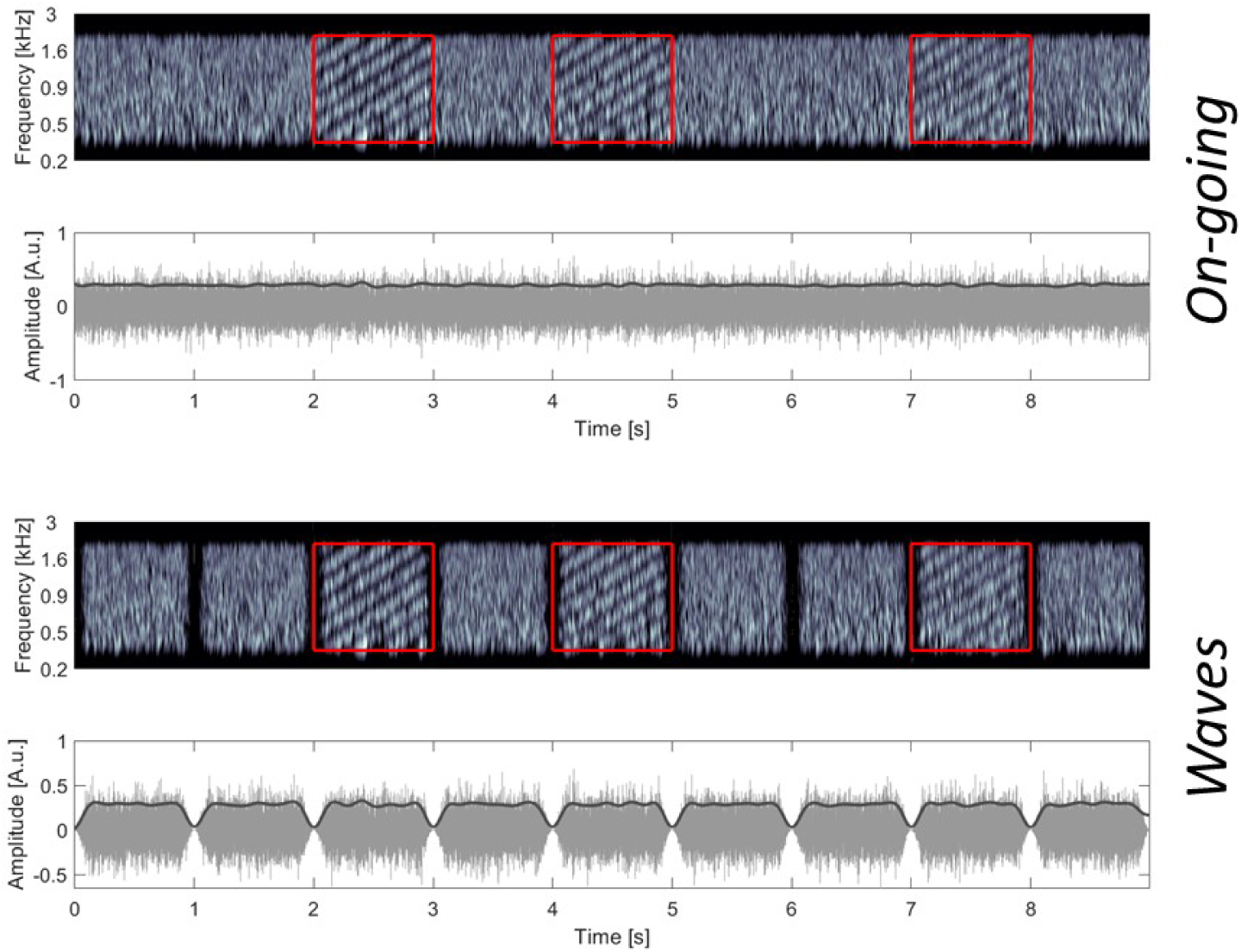
Stimulation paradigms. Top: *On-going* stimulation approach with running carrier noise and STM imposed in 1-s long time windows (red frames); Bottom: *Waves* approach with 1-s gated carrier noise and STM imposed in 1-s long time windows (red). In each panel, the upper plot shows a gammatone-filterbank-based auditory spectrogram with the red squares indicating the presence of an STM target (modulation levels from left to right: 0, −3, −6 dB FS); the lower plot shows the corresponding time signal in gray and its envelope in black.

**Table 1:**
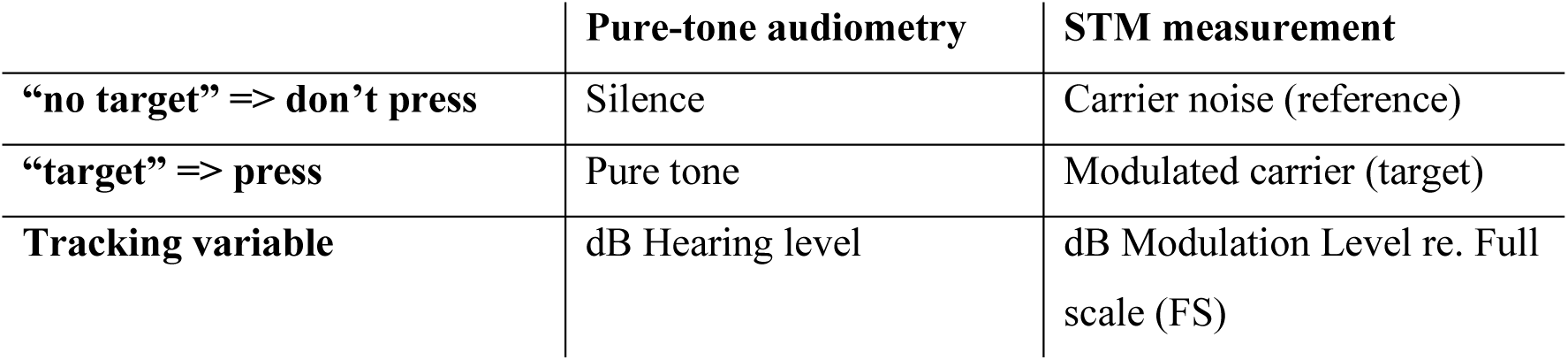
Comparison between pure-tone audiogram and proposed clinical STM measurement paradigm.

The threshold-seeking paradigm was designed to be as similar as possible to the Hughson-Westlake approach utilized in standard pure-tone audiometry (Carhart and Jerger, 1959). The tracking variable was the modulation level of the STM pattern, expressed in dB full scale (FS), i.e., relative to a full modulation being 20 log_10_ *m* |_*m*=1_ = 0 dB FS. Step sizes of 1.5 and 3 dB were used for both stimulation paradigms. The top panel of Figure 3 shows an example measurement conducted using a step size of 3 dB, according to the rules described in the following; note that the y-axis is reversed, such that an “ascending” run is plotted as moving downward and vice versa. Initially, the modulation level was set at its maximum of 0 dB FS. The examiner manually triggered when a target was to be played (i.e., when the STM was to be imposed on the running carrier noise). The test participants were asked to indicate that they had detected the presence of the STM by pressing a pushbutton. After a detection (“HIT”), the modulation level was decreased by 2 steps (here: 6 dB), making detection of the next target harder (descending run). After a failure to detect the target (“MISS”), the modulation level was increased by 1 step (here: 3 dB; ascending run), making detection of the next target easier, until there was again a HIT that triggered a descending run, and so forth. The modulation level of the detected STM signal at the end of each ascending run was considered a “threshold candidate” (trials 7, 10, 14, and 19 in the top panel of Figure 3). The procedure was terminated when three out of the last maximally five threshold candidates were identical (in Figure 3 only four candidates were needed to obtain three identical ones). The corresponding modulation level was termed the “stored threshold” (−9 dB FS in the example shown in Figure 3). A three-out-of-five Hughson-Westlake type procedure was preferred over the broadly accepted “two-out-of-three” approach commonly used in pure-tone audiometry (British Society of Audiology, 2018) because the response behavior of test participants in the STM detection task was found to be somewhat more variable as compared to pure-tone detection. The data from each run were then post-processed using a function fit to obtain a more fine-grained (fitted) threshold, as demonstrated in the bottom panel of Figure 3 and described in “Post processing”.

**Figure 3:**
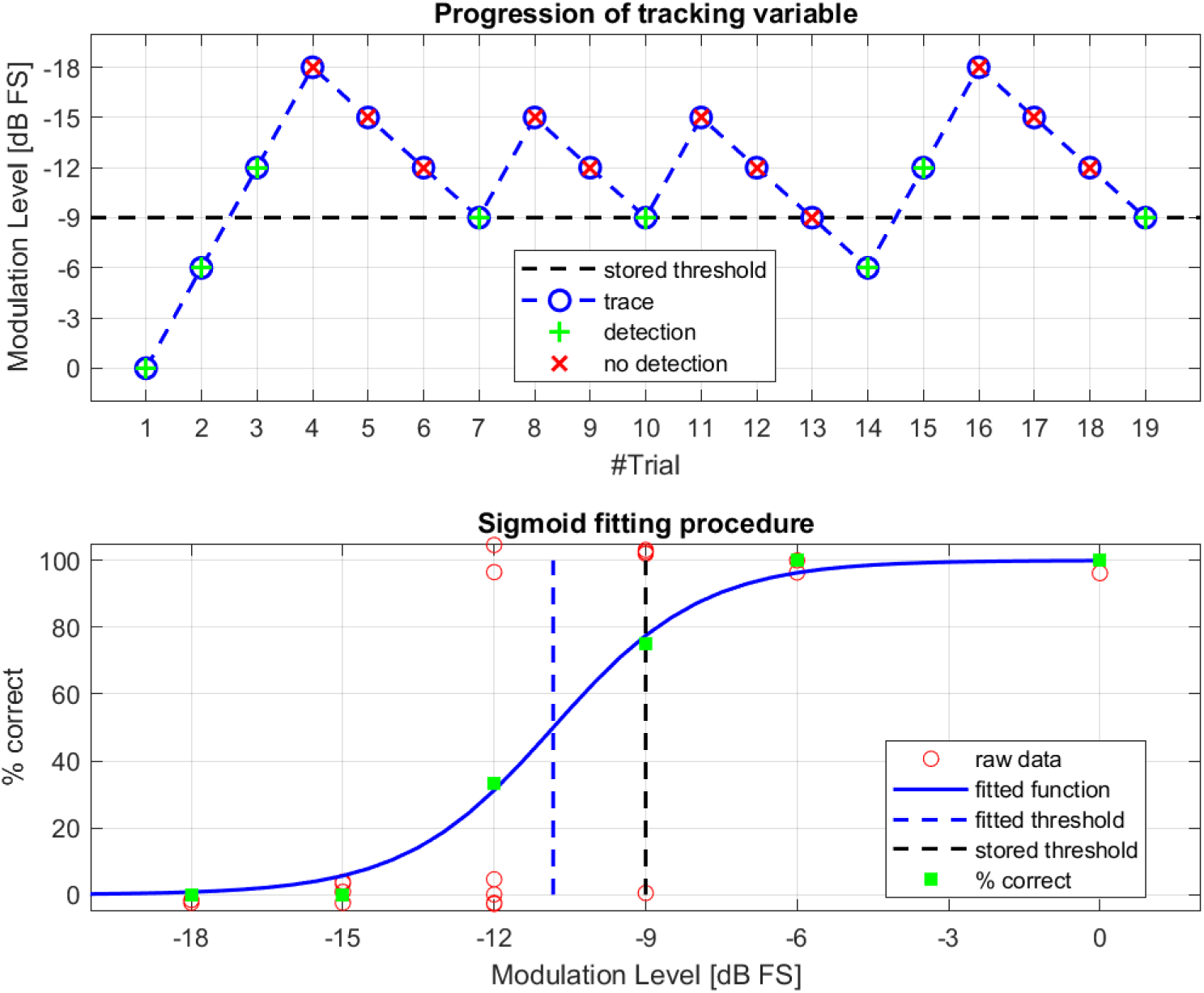
Measurement paradigm and data analysis approach. Top: example of measurement trajectory applying a Hughson-Westlake 3-out-of-5 criterion and using a step size of 3 dB; the modulation-level axis is reversed for consistency with study B and the classical audiometer hearing level display. Bottom: sigmoid function fitted to the data and corresponding 50% threshold. A small vertical jitter was added to the red markers representing the raw data.

### Hearing-loss compensation

To make sure that the stimuli were audible for all participants, an approach known as ‘sufficient audibility’ (Humes, 2007) was used (see also Zaar et al., 2023a, 2023b). The compensation was based on the ear-specific audiogram (see Figure 4), potentially leading to different hearing-loss compensation filters for the left and right ear.

**Figure 4:**
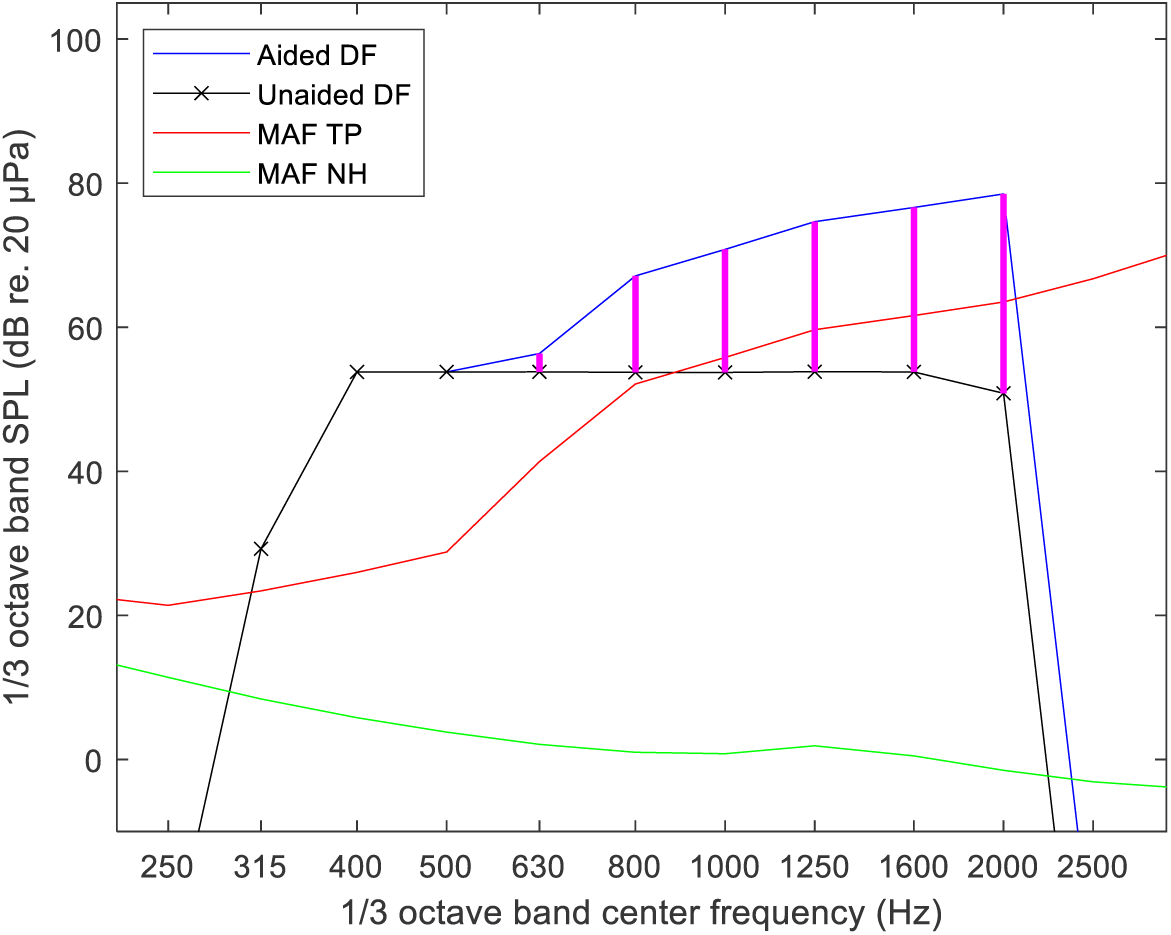
Hearing-loss compensation approach used to ensure audibility in the current study. The green line shows the minimum audible diffuse field for NH listeners (MAF NH). The red line shows the MAF for a HI test participant (MAF TP). The black line indicates the nominal diffuse-field (DF) levels of the stimulus. The blue line shows the amplified DF levels of the stimulus, ensuring at least 15 dB sensation level, accomplished by the gain depicted as magenta lines. All calculations were done per third-octave band.

Both the reference and target STM stimuli used in the experiment have the same long-term power spectral density, which was integrated into third-octave bands and compared with pure-tone hearing thresholds. According to Moore et al. (2008), third-octave bands are good approximations of the equivalent rectangular bandwidth of the auditory filters for people with mild-to-moderate hearing loss.

Starting from a nominal broadband stimulus level of 65 dB sound pressure level (SPL) in diffuse field (DF), the nominal stimulus levels were computed in third-octave frequency bands and compared with the ear-specific hearing-threshold levels interpolated from the pure-tone audiogram values onto the third-octave center frequencies. If the nominal stimulus level was less than 15 dB above the hearing threshold in any third-octave band within the stimulus bandwidth, the stimulus level in that band was amplified such that it was 15 dB above hearing threshold. The maximum output level in any third-octave band was limited to 90 dB SPL to avoid overstraining the Sennheiser (Wedemark, Germany) HDA200 headphones used in the experiment (limit determined by experimentation). The hearing-loss compensation procedure is illustrated in Figure 4, and is explained in detail below:

- The starting point is the minimum audible field for normal-hearing listeners (MAF NH, ISO 389-7, 2005, Table 1, col. 3), that is, the normal-hearing threshold for DF listening, expressed in third-octave band levels (green curve in Figure 4).
- The MAF NH curve is then translated into the (ear-specific) minimum audible field for the test participant (MAF TP) in question by adding the pure-tone audiogram, or HTL, values in dB to the MAF NH after interpolation onto the third-octave band center frequencies (red curve).
- This curve is then compared band by band to the nominal DF third-octave band levels of the stimulus (black curve).
- If the black curve is already 15 dB or more above the MAF TP, no gain is specified; if not, gain is specified such that the stimulus band power is 15 dB above the MAF TP (gain specification is indicated by the bold magenta lines and the resulting aided stimulus spectrum by the blue line in Figure 4).

The magenta lines thus constitute the gain specification for the hearing-loss compensation filter within the stimulus bandwidth covering the 1/3-octave band center frequencies from 400 Hz to 2000 Hz (both included). Below and above this frequency range, the gain specifications obtained for the 400-Hz and 2-kHz band were applied, respectively.

### Procedure and apparatus

All measurements were conducted in a single visit, which lasted between 1.5 and 2.5 hours. Audiograms measured within a year before the visit were used without re-measuring them unless the participant reported a change in their hearing ability. New audiograms were thus only measured for three of the 28 participants. Prior to the STM measurements, an otoscopy was performed to ensure that the ear canal was sufficiently free from ear wax.

Two measurement blocks were conducted, each with a given measurement paradigm (either *On-going* or *Waves*). Within each block, six measurement runs were performed: three consecutive runs with a step size of 1.5 dB, and three consecutive runs with a step size of 3 dB. The order of the blocks (i.e., measurement paradigms) and the order of the step sizes within each block were balanced across participants. Participants were required to take a 5-minute break between runs 1-3 and runs 4-6 within each block. In between blocks, a longer break (about 15 minutes) was scheduled.

Prior to each measurement block, test participants were instructed to listen to the “siren” sound in the noise and to press the pushbutton whenever they heard it. The participants were seated in an acoustically insulated listening booth wearing a pair of Sennheiser HDA200 headphones and a pushbutton (similar to those used in pure-tone audiometry) in hand. The examiner sat outside the booth operating a Windows-based PC running a Matlab (Mathworks, Nathick, Massachusetts, USA) software with a graphical user interface (GUI). The GUI allowed to start the measurement run, triggering continuous playback of the reference through an RME (Haimhausen, Germany) Fireface sound card and an SPL (Niederkrüchten, Deutschland) Phonitor mini headphone amplifier to the headphones worn by the test participant. The examiner could then manually control the modulation level and trigger playback of an STM target signal. The participant’s response was indicated on the GUI by a red light flashing. This was achieved by means of a custom-built battery-driven converter box, which sent a spike to the RME Fireface whenever the pushbutton was pressed. The headphones were equalized based on a measurement conducted using a Brüel & Kjær (Nærum, Denmark) hat and torso simulator (HATS type 4128-C), which was used as a good approximation of the sound transduction from the headphones to the individual participants’ eardrums. The stimulus level was set at a nominal level of 65 dB SPL in virtual free field, simulated by applying the diffuse-field-to-eardrum transformation from (Moore et al., 2008), amounting to 69.8 dB SPL at the HATS’ eardrum. The stimuli were presented to both ears simultaneously, with ear-specific hearing-loss compensation added where applicable according to the audiogram, as described in detail in the previous section.

A short training run was conducted at the beginning of each block. When a participant was initially unsure what to listen for, the examiner indicated visually when an STM target was being played during the training. The measurement runs were conducted according to the procedure described in “Stimulation and measurement paradigms”, starting at a modulation level of 0 dB FS and terminating the procedure when three out of the last maximally five threshold candidates matched. However, the target was presented three times at 0 dB FS before starting the adaptive procedure to ensure that the participant had a clear notion of what to listen for. In individual cases where a participant appeared to have lost the cue and suddenly started to perform much worse than before, the examiner started over from 0 dB FS during the run. Similar to pure-tone audiometry, the examiner was able to determine when an STM target was to be played such that arbitrary (“trigger-happy”) response behavior could be identified and counteracted by means of unpredictable target presentation times.

### Post processing

For each run, a stored threshold was obtained in the experiment, referring to the modulation level in dB FS at which three out of the last maximally five ascending runs ended, as described in detail in “Stimulation and measurement paradigms”. To obtain a more fine-grained threshold representation that is not quantized to the modulation-level grid defined by the step size, another threshold calculation method was considered as an alternative. This method utilized a sigmoid logistic function, which was fitted to the raw data collected during the measurement run. Although these raw data are binary as they simply consist of HIT/MISS for each STM target presentation, they can collectively be considered to follow a psychometric function, as shown in the bottom panel of Figure 3. The parameters *s*_50_ (slope parameter) and *x*_50_ (corresponding 50% point) of the function

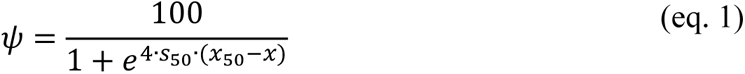

were found using a non-linear least-squares optimization on the raw data, with *ψ* representing the percent-correct score (100% or 0% in the raw data) and *x* denoting the modulation level. The midpoint *x*_50_ was termed the “fitted” threshold in dB FS. The slope *s*_50_ is given in *dB*^−1^ and is here always negative as the performance improves as a function of *x* (Brand and Kollmeier, 2002).

## Results and analysis

The STM detection thresholds reflect the modulation level necessary to just distinguish between a signal with the modulation present (target) and a signal without any imposed modulation (reference). Therefore, low thresholds indicate good performance and vice versa. To represent the thresholds in a visually intuitive fashion that represents performance directly (in analogy to the audiogram), the modulation-level axes have been reversed in the figures below, such that they show high values (indicating poor performance) on the left/bottom and low values (indicating good performance) on the right/top. A similar approach was also employed in study B, albeit using a modified definition of the tracking variable.

### STM detection group results

The four test conditions, i.e., the two measurement paradigms *On-going* and *Waves* combined with the two step sizes of 1.5 and 3 dB, were first compared at the group level. For this purpose, the mean value of the stored and fitted thresholds across the three runs conducted in each test condition was taken for each participant. It should be noted that every single run yielded a meaningful result according to the three-out-of-five measurement approach, albeit with a small number of runs where the examiner had to start over (i.e., go back to 0 dB FS) during the run.

Figure 5 shows that all test conditions led to quite similar stored and fitted thresholds on average. Overall, the stored thresholds spanned a range between −15 and −2.5 dB FS with a median at −9 dB FS, whereas the fitted thresholds spanned a range between −16.3 and −3.6 dB FS with a median of −10.6 dB FS. An analysis of variance (ANOVA) using a linear mixed-effects model with ‘test condition’ (*On-going*/*Waves* with 1.5/3 dB) and ‘threshold type’ (stored/fitted) as fixed effects and ‘participant’ as a random effect revealed that stored and fitted thresholds were significantly different (F(1, 27)=338.5, p<10^-16^), with the fitted threshold being 1.0 dB lower than the stored threshold on average. A much less pronounced yet just-significant effect was also found across test conditions (F(3, 81)=3.14, p=0.03).

**Figure 5:**
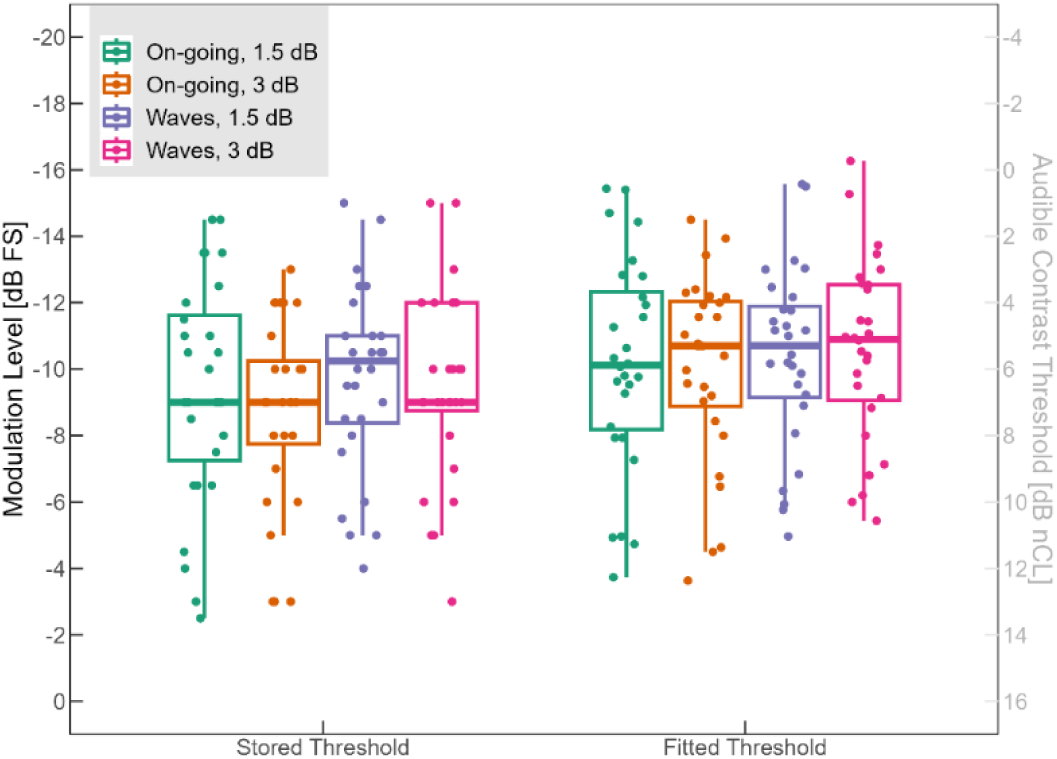
Boxplot of group data obtained in the STM experiment. Stored thresholds (left) and fitted thresholds (right) for the four test conditions consisting of two measurement paradigms and two step sizes. The points represent individual mean stored and fitted threshold values across the three runs. The boxplots show the median and the first and third quartiles (box), along with ±1.5 times the interquartile range (whiskers). The y-axis is reversed such that performance increases from bottom to top, with the left y-axis showing the modulation level and the right y-axis showing the normalized Contrast Level, introduced in Study B.

#### Measurement duration

Figure 6 shows the duration of individual measurement runs as a function of the measurement paradigms (left/right) and step sizes (red/blue). A clear trend can be observed: while the measurement paradigm only had a small influence, with *Waves* requiring slightly longer measurement time than *On-going* (112.9 s and 96.7 s, respectively), the step size had a strong influence, with 1.5 dB requiring a substantially longer measurement time than 3 dB (120.7 s and 88.9 s, respectively). There was a small but notable number of outliers, resulting from runs where the examiner had to start over multiple times due to the test participant losing focus or arbitrarily pressing the pushbutton. A two-way ANOVA using a linear mixed-effects model with ‘paradigm’ (*On-going*/*Waves*) and ‘step size’ (1.5/3 dB) as fixed effects and ‘participant’ as a random effect revealed that both the effect of paradigm (F(1, 27)=7.7, p<10^-2^) and the effect of step size (F(1, 27) = 31.8, p<10^-5^) on the measurement duration were statistically significant.

**Figure 6:**
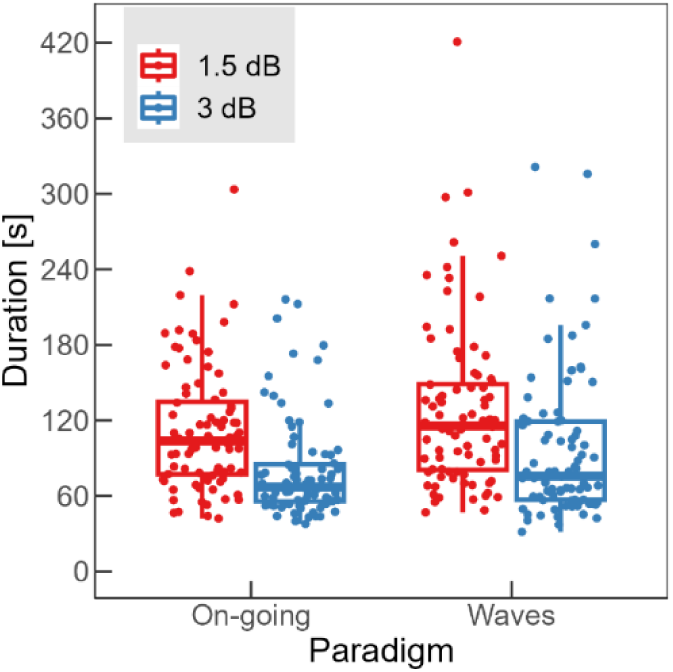
Boxplot showing the distribution of the duration of all measurement runs (points), as a function of stimulation paradigm (left/right) and for the two step sizes (red/blue). The boxplots show the median and the first and third quartiles (box), along with ±1.5 times the interquartile range (whiskers).

#### Test-retest reliability

To assess the test-retest reliability of the different combinations of measurement paradigm and step size, the thresholds collected across the three runs conducted in each test condition were compared. This was done separately for the stored and the fitted thresholds. For this purpose, the Intraclass Correlation Coefficient (ICC; Koo & Li, 2016) was computed for all possible pair-wise comparisons of the three runs conducted in each test condition, using a single-rating, absolute-agreement, two-way random effects model. Figure 7 depicts the results, with the gray and black symbols representing the ICCs obtained with the stored and fitted thresholds, respectively, and the vertical lines showing the respective 95-% confidence intervals. It can be observed that all ICCs indicate no less than “good” (0.75-0.9), some even “excellent” (0.9-1.0) reliability, with the fitted thresholds mostly leading to higher ICC values than the stored thresholds. The consistently highest ICCs were obtained for both measurement paradigms using the fitted thresholds and a step size of 1.5 dB.

**Figure 7:**
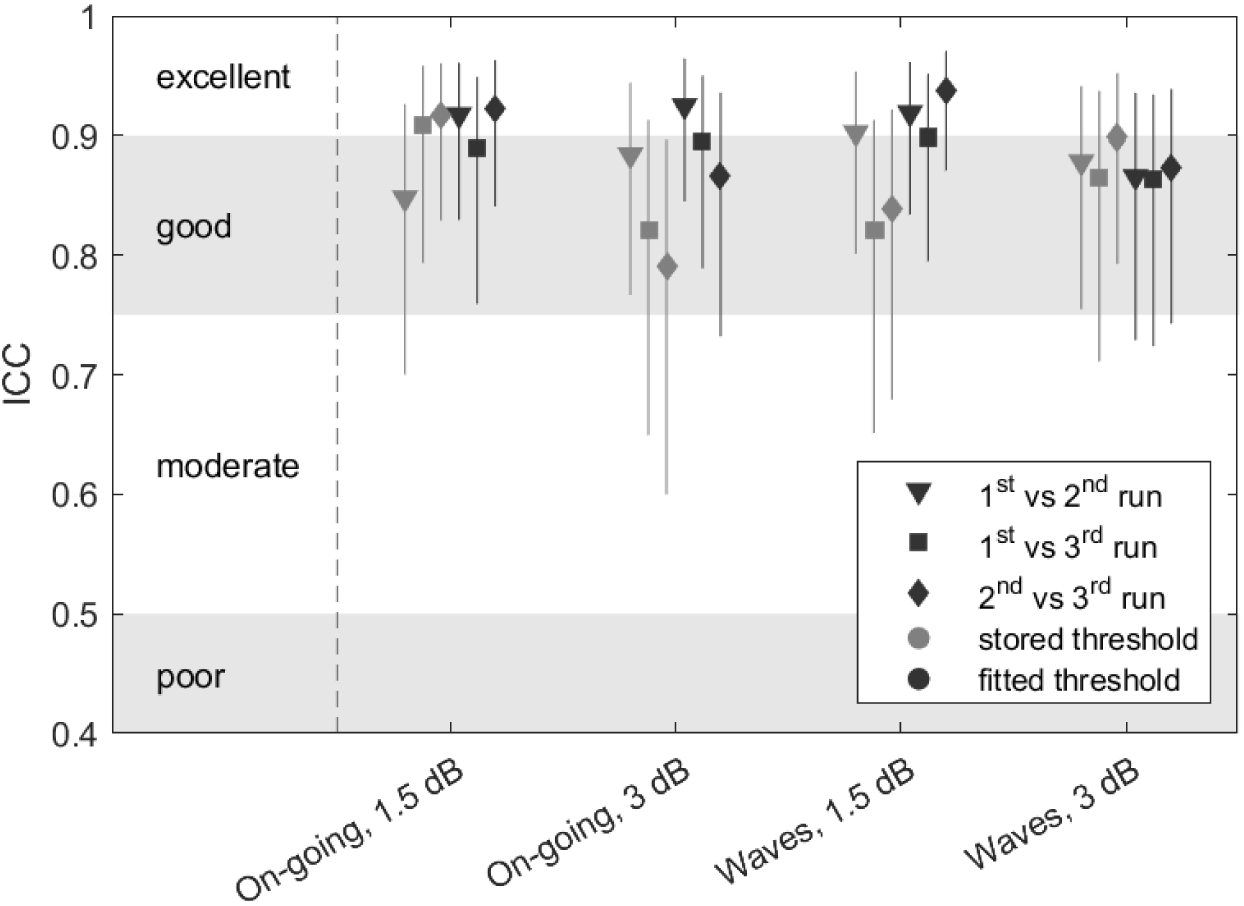
Intraclass Correlation Coefficient (ICC) calculated for all possible pair-wise comparisons of the three runs conducted in each test condition. The gray symbols indicate ICCs obtained with the stored thresholds while the black symbols denote ICCs obtained with the fitted thresholds. The vertical lines represent the 95-% confidence intervals.

#### Comparison with reference data from Zaar et al. (2023b)

The STM thresholds collected in the present study were compared to those measured about one year earlier for the same test participants with a 3-AFC paradigm by Zaar et al. (2023b). For this purpose, the mean value of the six thresholds collected by Zaar et al. (2023b) in test and retest – in the following termed the “reference threshold” – was compared to the mean value of the three thresholds collected in each of the four test conditions considered in the present study.

In a first step, the ICC was computed between the reference and the stored as well as the fitted thresholds for each of the four test conditions using a mean-rating, absolute-agreement, two-way random effects model. Figure 8 shows the reference thresholds as a function of the stored (left panel, gray symbols) and the fitted (right panel, black symbols) thresholds. Each dot represents a test participant, and the dashed lines show linear fits to the data. The respective test condition, ICC value, and the p-value corresponding to the ICC are shown in each subplot. The stored thresholds measured with the *On-going* paradigm showed only just-moderate agreement (ICC < 0.55, p > 0.04) with the reference thresholds, whereas the stored thresholds measured with the *Waves* paradigm showed an agreement at the upper end of the “moderate” category (ICC > 0.7, p < 0.04). For both measurement paradigms, the agreement was slightly better for the 1.5-dB than for the 3-dB step size (0.55 vs. 0.54, respectively, for *On-going* and 0.74 vs. 0.70 for *Waves*). In the right panel of Figure 8 it can be seen that the calculation of the fitted thresholds from the raw data improved the ICC, especially for the step size of 3 dB, such that the differences across step sizes became negligible. However, the fitted thresholds measured in the *On-going* paradigm still showed a weaker (moderate) agreement with the reference thresholds (ICC around 0.64, p < 0.02) than the fitted thresholds measured in the *Waves* paradigm (ICC around 0.78, p < 0.003; good agreement). It should be noted that the performance of some test participants may have been influenced by the presentation order of the test conditions in terms of losing focus due to fatigue. When factoring out the significant effects of presentation order in a multiple regression model (not shown here), the ICC for the *Waves* measurement paradigm even reached values of 0.82, indicating good agreement with the one-year-old reference thresholds.

**Figure 8:**
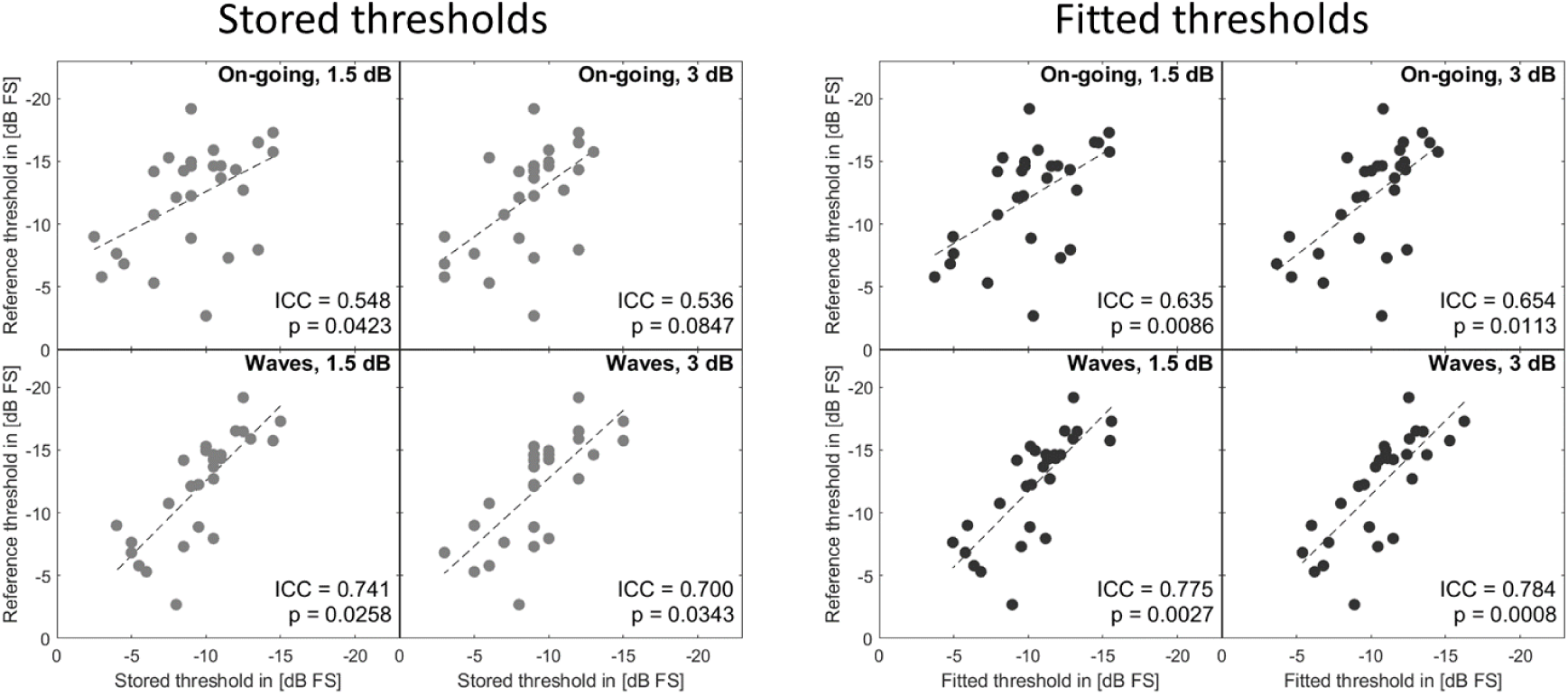
Reference STM thresholds measured by Zaar, et al. (2023b) as a function of the thresholds measured in study A with the two measurement paradigms and two step sizes. Left panel: stored thresholds; right panel: fitted thresholds. The respective test condition, ICC, and corresponding p-value are shown in each plot. The dashed lines represent linear fits to the data. The axes are reversed such that performance increases from left to right and from bottom to top.

In a second step, Bland-Altman plots (Bland and Altman, 1986) were obtained, comparing the fitted thresholds measured in the present study with the reference thresholds. Only the fitted thresholds were considered here as they proved to yield superior ICC results as compared to the stored thresholds. Figure 9 shows the difference between the fitted threshold m_Fitted_ and the reference threshold m_Ref_ as a function of their mean. The mean of the differences was around 2 dB for all test conditions, meaning that the measurement paradigms used in the present study were, on average, more challenging than the 3-AFC paradigm used in the reference study. The spread of the differences was somewhat smaller for the *Waves* paradigm (+/-2SD range of about 11 dB, bottom panels of Figure 9) than for the *On-going* paradigm (+/-2SD range of about 15 dB, top panels of Figure 9). In all test conditions, there was a clear downward trend, indicated by the dashed black regression lines. This trend implies that the better performers (found on the right side of each panel) showed a higher threshold (indicating worse performance and/or higher difficulty) in the present study as compared to the reference study, whereas the poorest performers (found on the left side of each panel) achieved about the same result in both studies.

**Figure 9:**
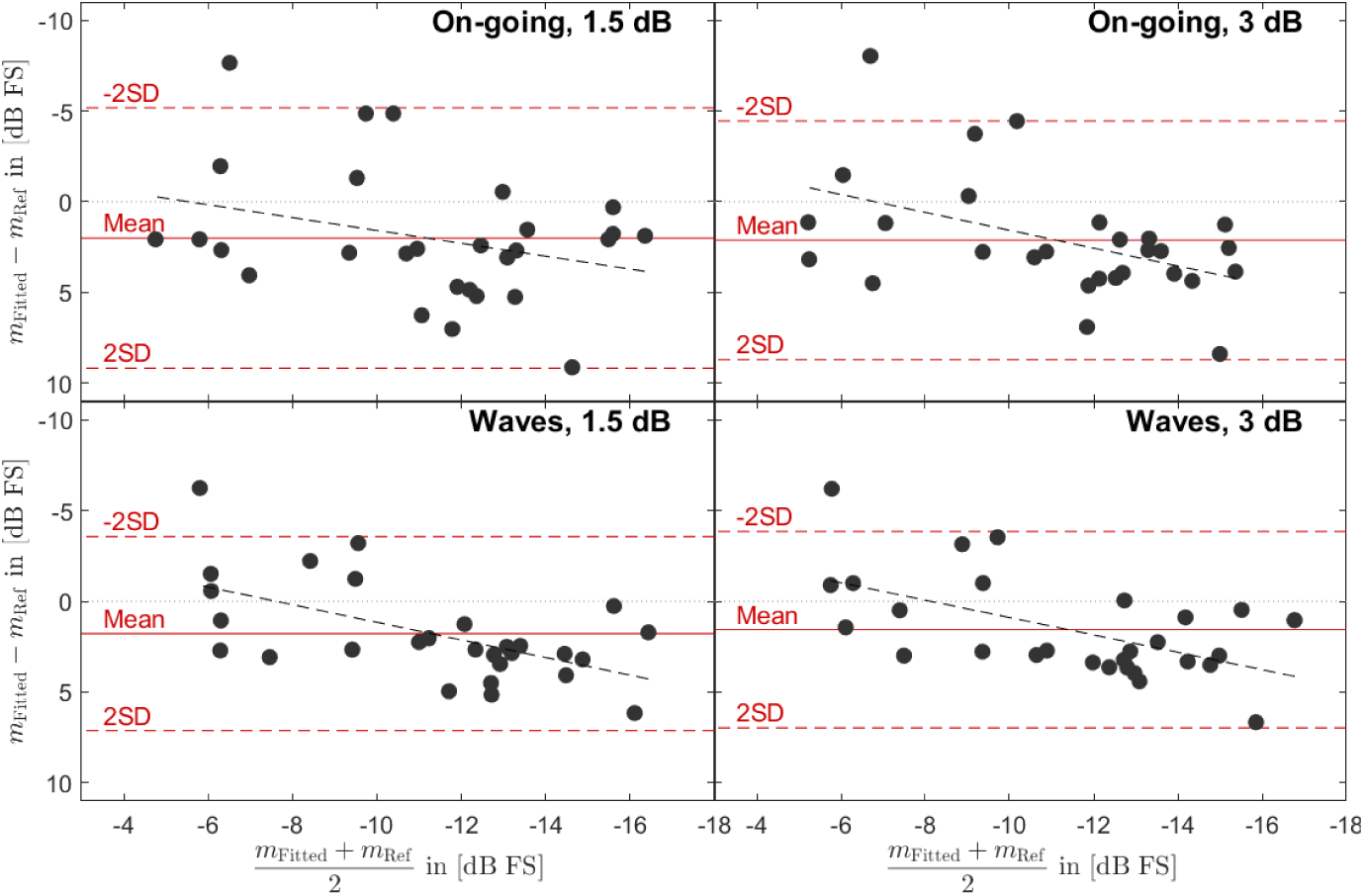
Bland-Altman plots comparing the fitted thresholds for the four test conditions with the reference thresholds. Each dot represents one test participant. The x-axis shows the mean between the fitted threshold *m*_*Fitted*_ and the reference threshold *m*_*Ref*_. The y-axis shows the difference between them. The solid red lines indicate the mean of the differences, the dashed red lines show the 95% confidence intervals. The dashed black lines show a linear fit to the data points. The axes are reversed, such that performance increases from left to right and relative performance in the present study as compared to the reference study increases from bottom to top.

## Summary and discussion

Study A proposed and investigated different approaches for clinical STM detection threshold measurements that can be conducted in a short amount of time with only a pair of headphones and a pushbutton. The two measurement paradigms *On-going* and *Waves*, combined with step sizes of 1.5 and 3 dB, were used to test 28 of the 30 HI participants that had already been tested with a 3-AFC paradigm by Zaar et al. (2023b) one year prior. Linear amplification was provided based on the audiogram where necessary to ensure at least 15 dB sensation level (SL). The basic stimulus design was similar to that used by Zaar et al. (2023b) and Bernstein et al. (2016), with a bandlimited noise carrier (354-2000 Hz) and upward-moving STM with 4 Hz and 2 c/o.

However, the carrier noise (reference) was here presented continuously, with the STM (target) imposed at the examiner’s request, such that the measurement procedure closely resembled the measurement method applied in pure-tone audiometry (with the carrier noise replacing the silence and STM imposed on the carrier noise replacing the pure tone). A strict three-out-of-five instead of the two-out-of-three criterion commonly used in pure-tone audiometry (British Society of Audiology, 2018) was employed, as the response behavior of test participants in the STM detection task is somewhat more variable as compared to pure-tone detection. Apart from the stored threshold resulting from the three-out-of-five criterion, a fitted threshold was also obtained from the data using a logistic function fit.

The following observations were made:

i. No substantial performance differences were found among the test conditions at the group level, but the fitted thresholds were found to be, on average, 1 dB lower than the stored thresholds (see Figure 5).
ii. The duration of a single run was short (roughly between one and two minutes), with the step size of 3 dB yielding significantly shorter duration than the step size of 1.5 dB (see Figure 6).
iii. The test-retest reliability was between “good” and “excellent” in all tested cases, with the step size of 1.5 dB yielding better reliability than the step size of 3 dB and with the fitted thresholds providing better reliability than the stored thresholds (see Figure 7).
iv. The agreement between the reference thresholds from Zaar et al. (2023b) and the stored thresholds was much better when using the smaller step size of 1.5 dB, but this difference disappeared when using the fitted thresholds, which globally yielded the far better agreement with the reference thresholds (see Figure 8). The agreement was globally substantially better for the *Waves* than the *On-going* paradigm (see Figure 8).
v. All measurement paradigms proposed here yielded overall higher thresholds than the 3-AFC paradigm, especially for the best performers (see Figure 9). This was likely a result of an increased difficulty due to fine-grained differences between segments in the (unmodulated) running carrier noise, as opposed to deterministic (“frozen”) carrier noise signals used in all three intervals of a given trial by Zaar et al. (2023b). However, a deterioration of the participants’ supra-threshold hearing over time (the two studies were conducted one year apart) could also explain the threshold elevation.

It is concluded from the above that the *Waves* paradigm should be preferred over the *On-going* paradigm and that the fitted thresholds should be preferred over the stored thresholds. When it comes to the step size, a trade-off between the measurement duration (shorter for 3 dB than for 1.5 dB) and the test-retest reliability (better for 1.5 dB than for 3 dB) was observed, meaning that a good compromise needs to be found.

Overall, the present study demonstrates that the 3-AFC STM detection test used in Zaar et al. (2023b) can be replaced with a much faster and clinically viable measurement paradigm, adding only a few minutes to the protocol of measurements in a clinical setting. It should be taken into account that a “good” agreement (according to ICC) between the reference data from Zaar et al. (2023b) and the preferred measurement paradigm from the present study represents an excellent result considering that the measurements were conducted one year apart. Furthermore, the differences observed between the reference STM test and the proposed clinical implementation mainly affected the better performers, which suggests that the clinical STM test can successfully detect reduced supra-threshold hearing abilities in a quick-and-simple test. Given the results reported by Zaar et al. (2023b), it is thus assumed that a clinical STM detection measurement paradigm based on the *Waves* approach with a suitable step size and thresholds re-calculated using a fitting function will yield results that are strongly related to ecologically valid speech outcomes in aided HI listeners and even predict the speech-intelligibility benefit induced by strong over moderate hearing-aid noise reduction.

## Study B: Clinical prototype of the Audible Contrast Threshold test and normative data for various transducers

### Background and test design

With a quick, simple, and thus clinically viable STM test the clinician will be able to determine how much “audible contrast” (described as modulation level in Study A) is required for an individual patient to detect the STM target. This will reflect the patients’ need for clearness to understand speech in noise, assuming audibility has been restored. This clinical STM test is named the Audible Contrast Threshold (ACT) test. The main aim of Study B was to refine and validate the approach suggested in Study A, and to assess the performance of young NH participants in the ACT test. These normative data are needed to evaluate whether a patient’s performance is within the normal range. In the same way that Reference Equivalent Threshold Sound Pressure Level (RETSPL) defines the zero-reference in pure-tone audiometry, a zero-reference for audible contrast perception is required for the test to assess supra-threshold hearing deficits. The secondary aims of Study B were to investigate the reliability of the test and to explore the possible effect of monaural vs. the default diotic binaural stimulus presentation. Furthermore, Study B included an evaluation of the potential differences among four different clinically relevant transducers. Finally, the data collected in Study B provides information on the time needed for a single ACT run.

The ACT test is based on the *Waves* approach recommended in Study A with fundamentally similar threshold-seeking procedure and stimuli specifications. To meet a compromise between measurement duration (larger step size preferred) and test-retest reliability (smaller step size preferred) a step size of 2 dB was chosen for Study B. Furthermore, the point of the psychometric function that corresponds to the fitted ACT was here established via analysis of the normative data. A number of additional design decisions were made to strengthen the clinical viability of the ACT test. Finally, a more clinically-friendly calibration procedure was introduced.

For the clinical ACT test the degree of modulation level is no longer described based on the physical properties of the stimulus (modulation level), but instead based on the auditory percept associated with it (audible contrast). Therefore, the threshold is defined as the normalized contrast level (nCL) in dB to provide a more clinically intuitive term, with 0 dB nCL being the zero-reference. Although the aim of the study was to establish this zero-reference in terms of contrast level, a pilot-data based estimate of the general ETML was set in the system prior the realization of the study to facilitate the consistency of the obtained thresholds (Westlake, 1943). The chosen relationship between modulation level in dB FS and nCL in dB nCL, is described in equation 2:

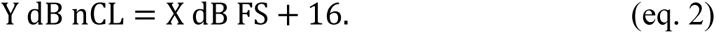

Therefore, full modulation level (corresponding to 0 dB FS in Study A) was transformed into 16 dB nCL (easiest), a modulation level of −16 dB FS in Study A was transformed into 0 dB nCL in Study B, and so forth. This has the advantage that the supra-threshold hearing deficit of the patient can be expressed in terms of “contrast loss” directly from the measured ACT. If a person has, e.g., an ACT of 8 dB nCL, the clinician can infer that the patient has an 8-dB contrast loss. During the implementation of the ACT test, the range of nCLs, the visualization of the results, and the adequate procedure to estimate the final threshold were discussed. Contrary to the STM test of Study A, the range of contrast levels is here limited between −4 to 16 dB nCL. This was based on an unpublished pilot study where NH listeners tended to be unreliable at very negative nCLs, and additionally motivated by the lack of clinical interest in differentiating between performance in the normal range and “super-performance” (similar to the audiogram conventions). Since the ACT test procedure resembles the pure-tone audiometry, the “tracking trace” (showing the progression of the tracking variable, analogous to the top panel of Figure 3) is presented in the clinical upside-down fashion, with −4 dB nCL at the top, in analogy to the audiogram. Finally, given that the “fitted” threshold was found to be more reliable than the “stored” threshold in Study A, it was decided to use a similar post-processing procedure to obtain the final ACT value. However, here, the data obtained in the sample of young NH listeners were used to establish an unbiased strong association between the stored and fitted thresholds. This was done because previous studies reported that the stored (Hughson-Westlake) threshold is well above the 50-% point (Marshall and Jesteadt, 1986) used in Study A.

Overall, the ACT test has been designed as a test of supra-threshold hearing abilities (specifically the audible contrast level), which is time-efficient, has similarities with an audiometry, and yields results that are comparable with previous STM tests.

## Methods

### Test participants

A total of 25 participants (females/males=14/11) participated in the study with inclusion criteria according to the ISO standard for preferred conditions for the determination of reference hearing threshold levels ISO 389-9 (2009). According to the standard, the age criterion for participation is 18-25 years on the day of testing and this was indeed the age range (mean: 23.3 years, standard deviation: 1.9 years) in this study. Another inclusion criterion was that test participants should have normal hearing and should be otologically normal, which was based on: a) otoscopy, b) tympanometry with a compliance peak within +/- 50 daPa of ambient pressure, c) the Annex A (normative) questionnaire for hearing tests ISO 389-9 (2009), and d) the pure-tone air-conduction audiogram measured at 0.125, 0.25, 0.5, 0.75, 1, 1.5, 2, 3, 4, 6, and 8 kHz. Specifically for this study, to guarantee no effect of hearing level on the performance in the ACT test, the maximum acceptable hearing level was set at 20 dB HL up to and including 6 kHz and 30 dB HL at 8 kHz. The participants’ audiograms are illustrated in Figure 10.

**Figure 10:**
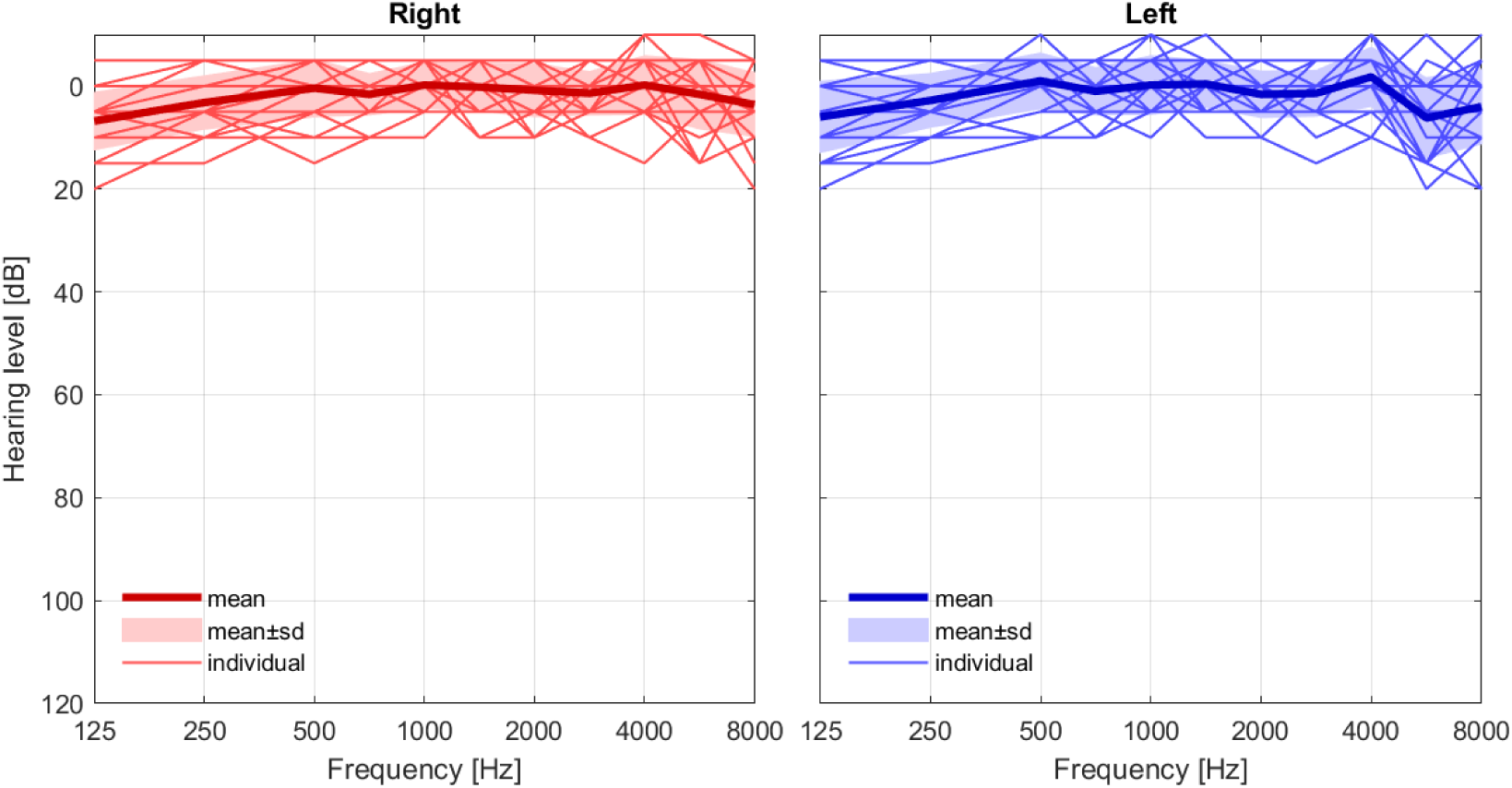
Pure-tone audiograms for 50 (25 left and 25 right) ears measured in study B with a RadioEar DD45 headset. The thin lines show the audiograms for individual participants, the thick lines the across-participant means, and the shaded areas reflect the across-participant standard deviations.

Test participants provided informed consent and the experiment was approved by the Science-Ethics Committee for the Capital Region of Denmark (Study B, reference H-1-2013-138). The test sessions lasted approximately one hour, and test participants were compensated with gift cards.

### Procedure and protocol

All test participants went through the preliminary investigations (otoscopy, questionnaire, tympanometry, audiometry) before conducting the ACT test to ensure fulfilment of the inclusion criteria. A standard procedure for measuring the audiogram was applied with the shortened version of the ascending method (two out three responses) and a step size of 5 dB, as described in ISO 8253-1 (2010). Each pure tone was presented manually for about 1 second, in response to which the test participant pressed a pushbutton when the stimulus was heard. For the audiometry and ACT measurements the test participant was positioned in a chair in a single-walled soundproof booth that met maximum permissible background noise as stated in (ISO 8253-1, 2010) and (ISO 8253-3, 2012). A window allowed the examiner to observe the test participant during data collection. The test participants were provided a short break between the audiometry and the ACT measurements.

The ACT values were collected using a 2-dB step size. The test participants were provided with both written-and oral instructions before the ACT test was started, see Text box 1 for the written instruction. The ACT test was started at 16 dB nCL, which is the easiest contrast level. The target was presented three times at 16 dB nCL to allow the listener to familiarize themselves with the target, whereafter the Hughson-Westlake procedure (Carhart and Jerger, 1959) followed (see “Stimulation and measurement paradigms” of Study A for further details). Special consideration is needed when the ceiling of −4 dB nCL is reached (the run is terminated when three out of the last five trials are correct responses at −4 dB nCL). Similar considerations apply at the 16-dB nCL floor level (albeit irrelevant for the present NH test participants). The final result – the fitted ACT – is determined by means of an estimated psychometric function to take into account all relevant responses of the listener. This is described in detail in the “ACT data post-processing” section below. The ACT value can vary between 16 dB nCL (worst performance) and −4 dB nCL (best performance). All ACT measurements were obtained with a clinical research prototype and administered by a trained audiologist.

#### Text box 1

##### Written instruction

*In this test, you will be wearing headphones on both ears and you will listen to a noise, which will be presented in waves. Every now and then the wave will be mixed with a siren-like sound. You can think of it as listening to the sound of the ocean, with the sound of a siren occurring now and then in the distance.*

*We would like to know how faint a ‘siren-sound’ you can just hear, when you listen for it. The strength of the siren-sound will therefore vary during the experiment.*

*Your task is to press the response-button when you hear the siren-sound.*

*The test will start at the easiest level, and gets then harder and harder.*

*Please ask if you have any questions.*

*Thank you very much for your participation!*

A total of six conditions combining different transducers and presentation modes (binaural/monaural) were considered. For the monaural condition, 13 participants had the left ear and 12 participants the right ear tested. The first measurement was always a binaural condition using the DD450 headset, whereafter the remaining five conditions were presented in a cyclic order with the second condition for each test participant advanced by one relative to the previous test participant (see Table 2). A short break was provided halfway through the six conditions to avoid fatigue as the ACT test requires a high level of concentration.

**Table 2:**
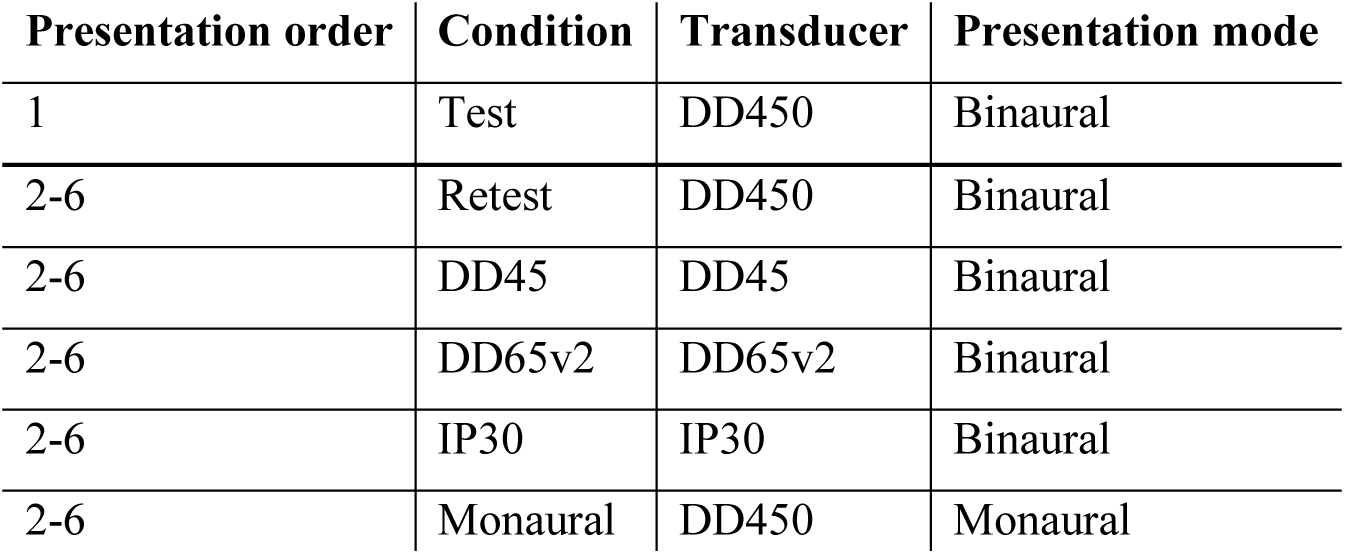
Overview of the six ACT measurements carried out in Study B with various transducers and presentation modes.

| Presentation order | Condition | Transducer | Presentation mode |
| --- | --- | --- | --- |
| 1 | Test | DD450 | Binaural |
| 2-6 | Retest | DD450 | Binaural |
| 2-6 | DD45 | DD45 | Binaural |
| 2-6 | DD65v2 | DD65v2 | Binaural |
| 2-6 | IP30 | IP30 | Binaural |
| 2-6 | Monaural | DD450 | Monaural |

#### Equipment

The Interacoustics (Middelfart, Denmark) AC40 Clinical Audiometer together with a RadioEar (Middelfart, Denmark) DD45 headset was used for measuring the pure-tone thresholds while the Interacoustics Titan was used for tympanometry. A research prototype software implemented in Matlab 2020a was used for collecting the ACT data, which was connected to a soundcard. The soundcard was further connected to a Lake People (Konstanz, Germany) G103-P headphone (HP) amplifier and a response box (similar to that used in study A).

As part of Study B, it was of interest to compare the ACT values obtained with different transducers. Therefore, we included the most common transducers used for audiometric tests (supra-aural, circumaural, and insert earphones), which differ in terms of bandwidth and dynamic range, see Table 3.

**Table 3:**
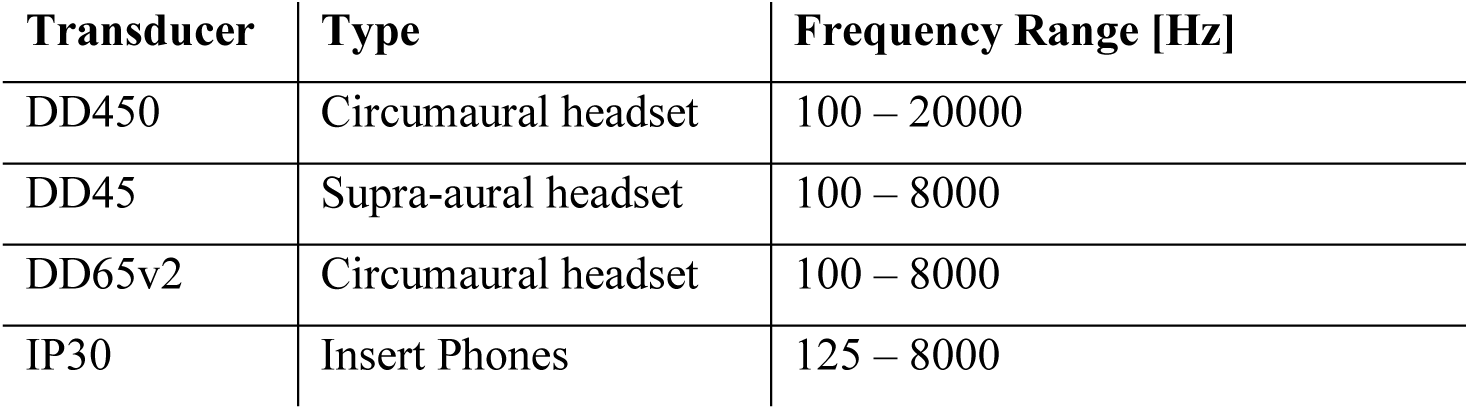
Overview of investigated transducers (all from RadioEar).

| Transducer | Type | Frequency Range [Hz] |
| --- | --- | --- |
| DD450 | Circumaural headset | 100 – 20000 |
| DD45 | Supra-aural headset | 100 – 8000 |
| DD65v2 | Circumaural headset | 100 – 8000 |
| IP30 | Insert Phones | 125 – 8000 |

#### Calibration

The main DD450 transducer was calibrated on a HATS manikin as in study A. Calibration filters for the other headsets were derived by considering differences between IEC 60318-1 (2009) Ear Simulator responses and RETSPL values for the DD450 versus the transducer in question (DD45 or DD65v2), so as to create the same (estimated) eardrum levels across frequency as with the DD450. The IP30 insert phones were equalized in a IEC 60318-4 (2010) Occluded Ear simulator and otherwise filtered in the same way as the DD450.

For future use, a calibration procedure based on standard measures is described in the supplementary material. The resulting differences in signal path gain were small compared with the above procedure: frequency response shapes were similar within ±1 dB, while the overall level was around 5 dB higher with the standard-measures procedure. For the NH test participants considered in the present Study B, this is considered inconsequential. For the HI test participants from Study A, the effect may potentially be larger, but in terms of the essential relative differences among test paradigms the results from Study A are nonetheless considered valid.

#### ACT data post-processing

To obtain a robust and accurate estimate of the ACT, two post-processing steps were conducted. In the first step, the “Threshold Candidate Window (TCW)” was extracted from the complete tracking trace of the trialed target contrast levels and the corresponding HIT/MISS participant responses. This was done to avoid undue influence from potentially poor performance in the beginning of a test run, which is not uncommon with inexperienced test participants. For normal test runs that concluded when the Hughson-Westlake criterion was fulfilled, the TCW included the last part of the tracking trace, starting from the last MISS before the fifth-to-last threshold candidate (see Figure 11). If the tracking trace only comprised four or three threshold candidates, the TCW was adjusted accordingly. For runs terminated due to the –4 dB nCL criterion, the TCW included the last five trials of the tracking trace.

**Figure 11:**
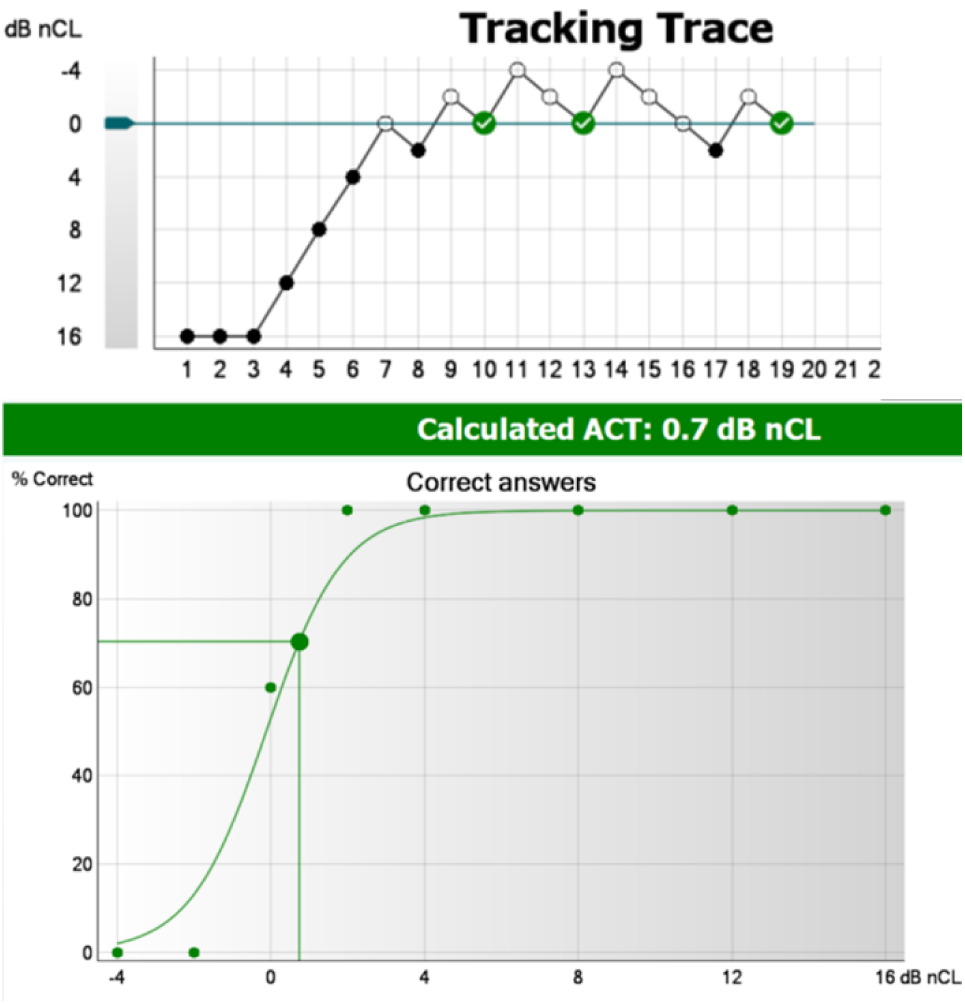
Example of a test run using the ACT research prototype. Top panel: Tracking trace, where the filled circles represent responses (HITs) and the open circles no responses (MISSes). The trials between 7 and 19 represent the threshold candidate window. Bottom panel: Psychometric function fitted to the data. The resulting fitted (calculated) ACT is indicated with the vertical line.

In the second post-processing step, the TCW was used to fit a psychometric function, inspired by the approach of (Rønne et al., 2017, 2013). To begin with, psychometric functions (for runs across all test participants and conditions that fulfilled the Hughson-Westlake criterion) were estimated with the slope *s*_50_ and the 50%-point *x*_50_ as free parameters (see eq. 1). To increase the robustness of the fitting procedure for individual runs, the median slope (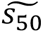 = 0.2518 *dB*^−1^) was selected and all psychometric functions were re-estimated using the fixed slope, 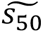, and the 50-% point as the only free parameter. Next, the target %-correct value, *ψ* = 70.22 %, was determined to minimize the mean difference between the ACT estimated via the psychometric function (at *ψ* % correct) and the stored threshold taken directly from the Hughson-Westlake criterion. This step eliminates the bias issue observed in Study A, where the stored thresholds were found to be consistently higher than the fitted thresholds. Thus, the final result of an ACT test (i.e., the participant’s ACT) is found by estimating a psychometric function from the TCW using the fixed slope, 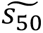, and determining the contrast level corresponding to the target %-correct value *ψ*.

## Results and analysis

Figure 12 depicts the ACT values of the 25 participants in the six measurement conditions. The median ACT values in the main DD450 test condition is very close to 0 dB nCL, indicating that the predicted zero-reference for the dB nCL scale from eq. 2 was indeed correct. Further, the results from the DD450 test condition indicate a range of normal performance from −4 to 4 dB nCL. Otherwise, Figure 12 indicates only small differences among the six test conditions; detailed analyses are presented below.

**Figure 12:**
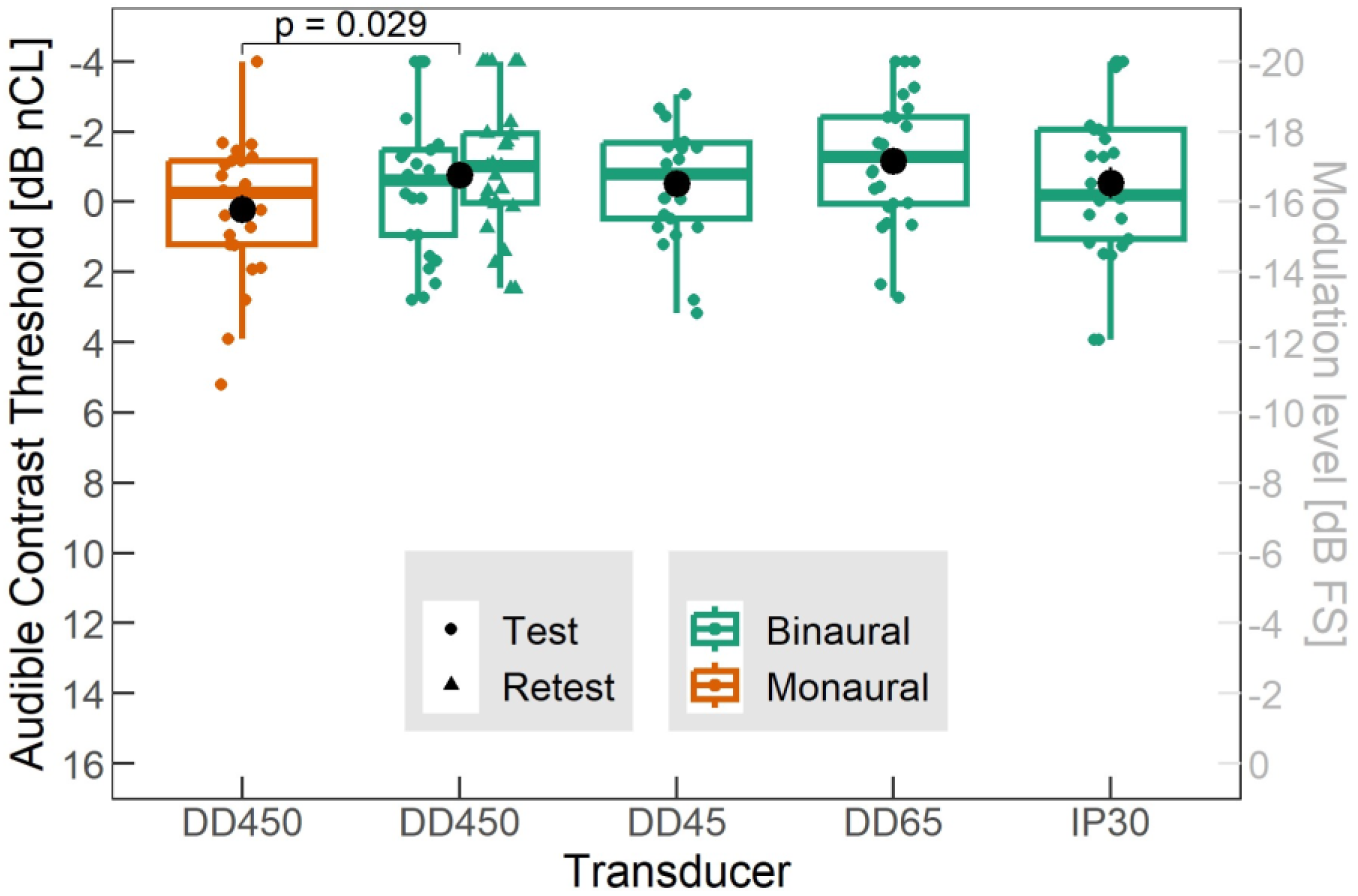
Audible contrast thresholds in dB nCL (left y-axis) from 25 test participants in six test conditions, as indicated. The black dots represent mean values. The boxplots show the median and the first and third quartiles (box) along with ±1.5 times the interquartile range (whiskers). The right-hand y-axis indicates the corresponding modulation level in dB FS, as used in Study A.

### Test-retest reliability

Test-retest reliability was assessed in two steps by comparing the DD450 test condition (always measured first) and the DD450 retest (measured as condition 2-6), first by the Bland and Altman method (Bland and Altman, 1986) and secondly with a paired t-test. Normality of the distribution of differences between the two test runs was confirmed by a Shapiro test (Royston, 1982), p = 0.39. Figure 13 presents a Bland-Altman plot of the test-retest data (Bland and Altman, 1986), which reveals a mean difference between test and retest of 0.5 dB. However, a paired t-test (t(24) = 1.1, p = 0.29) indicated no significant difference between the two. Figure 13 additionally suggests no systematic pattern in the test-retest differences. The standard deviation (SD) on a single measurement was found to be 1.5 dB, i.e., lower than one step size (2 dB). The limits of agreement (the mean +/- 2SD) are 3.5 and −2.5 dB, as illustrated in Figure 13.

**Figure 13:**
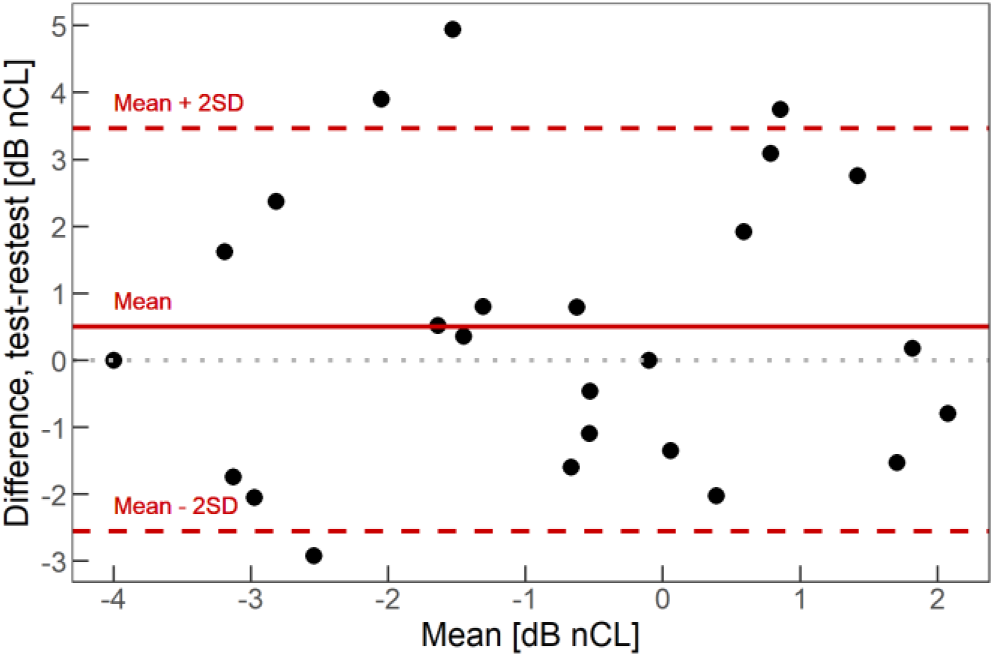
Bland-Altman plot showing the test-retest differences vs. the means of test and retest results. The red solid red line (mean) and dotted red lines (mean +/- 2SD) illustrate, respectively, the bias of 0.5 dB and the limits of agreement which are 3.5 and −2.5 dB. Each dot represents data from an individual participant.

### Monaural versus binaural presentation

As the test-retest difference was found not to be significantly different from 0, the means of the DD450 test and retest runs were used to investigate the effect of monaural vs. binaural stimuli presentation. A paired t-test revealed a significant difference between monaural and binaural stimuli presentation with a mean difference of 1.0 dB (t(24) = 2.3, p = 0.029), indicating that performance was 1 dB better (lower ACT) for binaural presentation as compared to monaural presentation. This is illustrated in Figure 12.

#### Effect of transducer and presentation order

To investigate whether the ACT values had been significantly affected by the different transducers or the presentation order, we performed an ANOVA using a linear mixed-effects model including all the binaural conditions outcomes, with ‘transducer’ and ‘order’ as fixed effects and ‘participant’ as a random effect. To improve the power of the model, both DD450 test and retest measurements were included, as they were not significantly different. The results indicated that neither the main effects of transducer and order, nor their interaction, were significant (transducer: F(3, 102.4) = 0.05, p = 0.98; order: F(1), 97.5) = 0.32, p = 0.57; interaction: F(3, 103.4) = 0.29, p = 0.83). Furthermore, using a backwards stepwise reduction of the model resulted in a model with no factors (null-hypothesis model), confirming that neither the transducer nor the presentation order showed a significant effect on the ACT.

### Measurement duration

A histogram of the measurement durations for all conducted measurement runs is presented in Figure 14. The mean test duration across all listeners and all test runs was 1 minute and 15 seconds (75.5 seconds, SD=23.8) with 75% of the listeners completing the test in less than 1.5 minutes (89 seconds) and the longest test run lasting 2.5 minutes (148 seconds).

**Figure 14:**
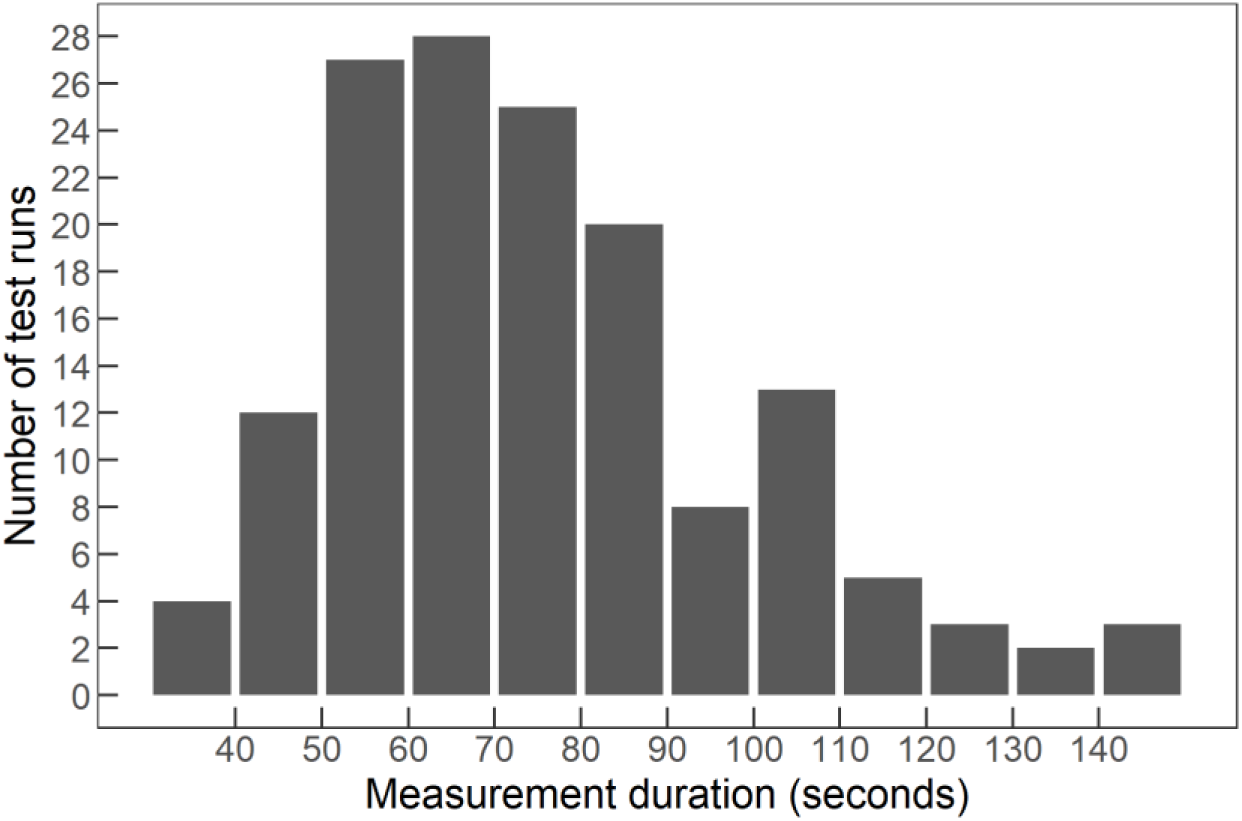
Histogram of measurement duration for all 25 test participants and all six conditions (i.e., a total of 150 runs).

## Summary and discussion

Study B investigated the ACT-test performance for 25 young NH listeners, who were assumed to have normal contrast thresholds. The test-retest reliability, the effect of binaural vs. monaural stimulation, as well as the effect of four different transducers were investigated while the measurement duration was tracked to assess clinical viability. The presentation order for the six measurement runs varied for each test participant (except the binaural/DD450 main test condition which was always presented first) to balance out effects of training and/or fatigue. As an ANOVA did not show any significant effect of presentation order, training and/or fatigue were not found to affect the results significantly. Specifically, the difference between the main DD450 test and retest conditions was found not to be significant.

The following observations were made:

i. With a test SD of 1.5 dB and no significant effect of training, clinicians can expect limited variation in the ACT value, and it is thus suggested that one test run is sufficient to obtain a reliable threshold. Attention should be paid to whether the patient understood the instruction or presses the response button unexpectedly often; if in doubt about the behavior or the result, it is advised to repeat the test.
ii. The monaural condition was found to show significantly higher (poorer) thresholds as compared to the binaural condition with an average difference of 1 dB (see Figure 12).
iii. The analysis of the results for four different transducers showed no effect of choosing one transducer over the other, and the examined transducers can therefore be applied interchangeably (see Figure 12).
iv. All measurement runs showed a mean duration of 1 minute and 15 seconds, which bodes well for the clinical viability of ACT as it may be considered clinically acceptable (see Figure 14).
v. Study B validated the approach suggested in Study A with a step size of 2 dB and defined the performance of young NH participants in the ACT test to be between −4 to 4 dB nCL measured binaurally with either a DD450, DD45, DD65 or IP30 transducer (the poorest performance observed in any of the binaural conditions was 3.9 dB nCL). If ACT is measured monaurally, the normal contrast level performance is defined as ranging from −4 to 5.5 dB nCL (the poorest performance observed in the monaural condition was 5.2 dB nCL).

For future directions, the test-retest variability should be investigated with the ICC including data from a clinical population with a wide variety of ACT values. Furthermore, it should be considered to further establish the range for a mild, moderate, and severe contrast loss, analogous to the division of the audiogram into mild, moderate, and severe hearing (sensitivity) loss. In this study, four different transducers, commonly used in hearing care, were considered for the investigation. However, for future application, it could be considered to present the stimuli directly from a hearing aid and/or from a smartphone application. Such alternative presentation modes would of course require careful consideration regarding calibration.

## Overall summary and conclusions

The present study introduces the ACT test, an STM detection test with built-in audibility compensation that was specifically optimized for clinical use. The ACT test can be conducted with standard clinical audiometry equipment (pushbutton and headphones) and administered by an audiologist using a measurement procedure that very strongly resembles the approach used in pure-tone audiometry. The present study consists of two sub-studies: in Study A, the stimulation, measurement, and data-analysis paradigms were developed and validated in hearing-impaired (HI) listeners; in study B, the findings from study A were used to refine the approach and normative data were collected in young normal-hearing listeners, along with an assessment of different transducers and presentation modes.

Study A tested two different stimulation paradigms, one with continuous noise (*On-going*) and one with windowed noise (*Waves*). The basic stimulus design was based on literature (Bernstein et al., 2016; Zaar et al., 2023a, 2023b), with a bandlimited noise carrier (354-2000 Hz) and upward-moving STM with 4 Hz and 2 c/o. The carrier noise (reference) was presented continuously, with the modulation (target) imposed at the examiner’s request, such that the measurement procedure closely resembled the measurement method applied in pure-tone audiometry (with the carrier noise replacing the silence and modulation imposed on the carrier noise replacing the pure tone). The two measurement paradigms *On-going* and *Waves*, combined with step sizes of 1.5 and 3 dB, were used to test 28 HI participants for whom reference data from a 3-alternative forced-choice (3-AFC) high-precision spectro-temporal modulation detection test were available (Zaar et al., 2023b). The two paradigms were administered using a Hughson-Westlake tracking procedure with a three-out-of-five criterion. A logistic function fit was applied to the collected data to refine the threshold estimate. Based on an assessment of the test duration, test-retest reliability, and agreement with the 3-AFC reference data from Zaar et al. (2023b), Study A concluded that (i) the *Waves* paradigm was superior to the *On-going* paradigm, (ii) the logistic-function based refinement of the thresholds was meaningful, and (iii) the choice of step size was a compromise between measurement duration (shorter for 3 dB than for 1.5 dB) and test-retest reliability (better for 1.5 dB than for 3 dB).

In the light of the findings from Study A, Study B employed the *Waves* stimulation paradigm with a step size of 2 dB and a logistic function-fitting procedure that was refined as compared to the approach used in Study A. Twenty-five young normal-hearing listeners were tested using four transducers commonly used in clinical practice and a monaural (in addition to the standard binaural) presentation mode. The results indicated (i) no effect of the different transducers on performance, (ii) a good test-retest reliability of the test, (iii) a slightly but significantly reduced performance (1 dB higher thresholds) for the monaural presentation mode, and (iv) an average test duration of 1 minute and 15 seconds. The data obtained in the standard condition established a normative baseline for performance in the test, which served to transform the physical modulation level (in dB Full Scale) into the normalized Contrast Level scale (given in dB nCL), with 0±4 dB nCL indicating normal contrast sensitivity and values greater than 4 dB nCL (up to a maximum of 16 dB nCL) indicating a contrast loss. The nCL scale has been conceived in analogy to the Hearing Level scale, which is a normative representation of pure-tone sensitivity in dB HL (as opposed to the physical dB sound pressure level).

Overall, the results indicate that the ACT test may be considered a reliable, quick-and-simple (and thus clinically viable) test of STM sensitivity. The ACT can be measured directly after the audiogram using the same technical set up, adding only a few minutes to the process. Given the strong connection between STM sensitivity and supra-threshold speech-in-noise reception (e.g., with hearing-aid amplification), ACT yields a highly useful clinical diagnostic that is complementary to the audiogram. More research is needed to establish meaningful categories of contrast loss (mild, moderate, etc.), to fully validate the test in a clinical setting, and to confirm its predictive power with respect to speech-in-noise reception.

## Data Availability

All data produced in the present study are available upon reasonable request to the authors

## Acknowledgement

The authors would like to thank all test participants who took part in Study A and Study B. Furthermore, we would like to thank Jaime Undurraga for his support with statistical analysis and figure plotting for Study B, as well as for his helpful comments on an earlier version of the manuscript. Furthermore, we would like to acknowledge James Harte, Thomas Behrens, Thomas Lunner, and Bue Kristensen for their support of the study. This work was partially supported by the Innovation Fund Denmark (9090-00089B and 1044-00071B) and the William Demant Foundation (case number 20-2448).

## Abbreviations

ANOVA: Analysis of Variance
ACT: Audible Contrast Threshold
c/o: Cycles/Octave
dB nCL: Decibel normalized Contrast Level
DF: Diffuse Field
ETML: Equivalent Threshold Modulation Level
GUI: Graphical User Interface
HI: Hearing-Impaired
HL: Hearing Loss
ICC: Intraclass Correlation Coefficient
LME: Linear Mixed Effect
MAF NH: Minimum Audible Field for Normal-Hearing listeners
MAF TP: Minimum Audible Field for Test Participant
FS: Full Scale
NH: Normal-Hearing
RETSPL: Reference Equivalent Threshold Sound Pressure Level
SL: Sensation Level
SPL: Sound Pressure Level
STM: Spectro Temporal Modulation
SRTs: Speech Reception Thresholds
SD: Standard Deviation
TCW: Threshold Candidate Window
2-AFC: Two-Alternative Forced-Choice
3-AFC: Three-Alternative Forced-Choice

## Supplementary material: ACT clinical calibration procedure

To ensure that the ACT stimuli are reproduced with the intended frequency-specific sound pressure level, a three-step calibration procedure is employed. In step 1, the frequency response of the used transducer is equalized on a reference coupler, including the soundcard’s digital-to-analog converter and the headphone amplifier. For the transducers used in the present study, the DD450, DD65v2, and DD45 headsets were measured on an IEC 60318-1 (2009) Ear Simulator (with the “flat-plate coupler” for the circumaural headsets), while the IP30 insert phones were measured in an IEC 60318-4 (2010) Occluded Ear simulator (“711 coupler”). Designing and adding to the signal path a filter to invert the coupler frequency response (in the frequency range of interest) essentially forms a transducer with unit-gain response from digital signal value to sound pressure in Pascal (Pa), when measured on the coupler. In step 2, the coupling to real ears is considered by transducer and coupler-specific RETSPL values (ISO 389-1, 2017). By adding to the signal path a filter with a magnitude response matching the transducer-specific RETSPL values, a digital pure tone with an RMS level of 0 dB (with reference to 20‧10^−6^ “digital Pa”) will correspond to the average hearing threshold, or 0 dB HL. The final step 3 transforms the ACT stimuli, which are digitally scaled to their nominal SPL (diffuse field), to dB HL. This is achieved by adding to the signal path a filter with a magnitude response matching the negative (in dB) MAF (Minimum Audible diffuse Field, ISO 389-7, 2005). In practice, the transducer linearization (step 1 above) is accomplished with a transducer-type-specific IIR filter designed from the median response among typically 10 measured transducers. The IIR filter order varies with transducer type. Step 2 and 3, together with the hearing-loss compensation described above, are combined into a single FIR filter with 512 coefficients. To accommodate asymmetrical hearing-loss configurations, separate FIR filters for the left and right ear audio channels are created.

Finally, the nominal diffuse field ACT stimulus level is set to 62.5 dB SPL, which is the same as the level of the International Speech Test Signal (ISTS, (Holube et al., 2010)) within the ACT stimulus bandwidth (354 – 2000 Hz).

## Notes

### Competing Interest Statement

All authors are employed by Demant A/S.

### Funding Statement

This study was partially funded by the Innovation Fund Denmark (9090-00089B and 1044-00071B) and the William Demant Foundation (case number 20-2448).

### Author Declarations

Study A was approved by the Science-Ethics Committee for the Capital Region of Denmark (reference H-16036391). Study B was approved by the Science-Ethics Committee for the Capital Region of Denmark (reference H-1-2013-138).

